# The Correlation Between Serum Glucose-Potassium Ratio and Mortality Risk in Sepsis Patients: Analysis based on the MIMIC-IV database

**DOI:** 10.1101/2025.05.26.25328362

**Authors:** Junhui Kang, Yonghao Yu

## Abstract

**Background:** Sepsis is a systemic inflammatory response syndrome triggered by infection, often leading to multiple organ dysfunction and high mortality rates. Despite advancements in critical care, predicting the prognosis of sepsis remains challenging. This study explores the predictive value of the Glucose-Potassium Ratio (GPR) for all-cause mortality (ACM) in sepsis patients.

**Methods:** This retrospective cohort study utilized data from the MIMIC-IV 3.0 database, which included adult sepsis patients admitted to the ICU between 2008 and 2022. A total of 9,360 patients were analyzed based on GPR stratification (Q1 ≤ 25.35, Q2 = 25.35–34.69, Q3 ≥ 34.69). Kaplan-Meier survival curves, Cox regression models, and restricted cubic splines (RCS) were employed to assess GPR’s association with mortality at 28 days, 90 days, 180 days, and 1 year. Subgroup analyses based on age, gender, race, and comorbidities were also performed.

**Results:** Higher GPR was associated with significantly reduced mortality at all time points (28 days, 90 days, 180 days, and 1 year), with hazard ratios for Q3 ranging from 0.71 to 0.79 (P < 0.001). Kaplan-Meier curves showed better survival for patients with higher GPR. Subgroup analysis revealed stronger protective effects in older adults (≥60 years), females, and Asians. GPR outperformed glucose and potassium alone in predicting mortality, with higher AUC values for GPR across different time points.

**Conclusion:** GPR is an independent predictor of sepsis mortality, with a threshold of ≥34.69 associated with reduced risks of mortality at multiple time points. It offers a superior predictive capacity compared to glucose or potassium levels alone, supporting its potential use as a clinical tool for early risk stratification and individualized treatment of sepsis patients.

## Introduction

Sepsis, a life-threatening organ dysfunction resulting from a dysregulated host response to infection[1], involves complex pathophysiological and biochemical abnormalities. Along with septic shock, it remains a critical global health challenge, affecting millions of people annually[2]. Recent epidemiological data reveal a concerning global sepsis burden, with 48.9 million cases and a 20% mortality rate[3]. In China, the incidence of hospitalization has steadily increased, from 328.25 (95% CI 315.41-341.09) to 421.85 (95% CI 406.65-437.05) cases per 100,000 people between 2017 and 2019, placing a significant strain on both healthcare systems and the economy[4]. Despite progress in management strategies, accurately predicting prognosis remains a challenge due to the multifactorial pathophysiology of sepsis, which includes multi-organ dysfunction[5].

This underscores the urgent need for improved early recognition and intervention. Current prognostic reliance on clinical scoring systems (SOFA, qSOFA, APACHE II)[1,6,7] faces limitations in both timeliness and precision. While these tools offer preliminary assessments of severity, they often fail to account for dynamic pathophysiological changes that are crucial for accurate risk stratification. This diagnostic gap has driven the search for novel biomarkers with greater sensitivity and clinical utility.

Emerging evidence underscores the prognostic significance of metabolic biomarkers, particularly serum glucose and potassium levels. Stress-induced hyperglycemia, mediated by neuroendocrine activation and immune dysregulation[8,9], has been consistently linked to poor outcomes in critical conditions, including intracranial hemorrhage[10,11]. Interestingly, while hypokalemia shows no significant correlation with outcomes in hemorrhagic stroke[12], the glucose-potassium ratio (GPR) demonstrates unique prognostic value due to their interdependent metabolic relationships[13,14]. Preliminary studies in neurological injuries, such as carbon monoxide poisoning, suggest GPR’s potential as a severity marker[15], although its role in sepsis remains underexplored.

Notably, large-scale investigations into the predictive capacity of GPR in sepsis are lacking. This study thus utilizes the MIMIC-IV database to systematically evaluate GPR’s association with sepsis mortality and its potential as an independent prognostic indicator. Through a comprehensive analysis of GPR’s clinical relevance, we aim to establish an evidence base for optimizing early detection, risk stratification, and personalized therapeutic strategies in sepsis management.

## Methods

### Data Sources

The Medical Information Mart for Intensive Care (MIMIC-IV v3.0) database, developed collaboratively by Beth Israel Deaconess Medical Center (BIDMC), the MIT Computational Physiology Laboratory, and Philips Healthcare, is a publicly accessible repository containing de-identified critical care data from 364,627 hospital admissions between 2008 and 2022[16]. This third-generation database improves upon its predecessor, MIMIC-II, through structural optimization, expanded clinical variables, and enhanced data accessibility. As a retrospective analysis of anonymized records, this study was exempt from institutional review board approval and waived the requirement for informed consent under 45 CFR 46.104(d)(4)[17]. The principal investigator completed CITI program certification (ID: 66195794) to ensure ethical data handling. Study Population

As shown in Fig. 1, using the Sepsis-3 criteria[1], we initially identified 95,458 septic patients requiring ICU admission. A rigorous exclusion protocol was applied: 1) missing initial ICU glucose/potassium measurements; 2) age <18 years; 3) ICU length of stay <24 hours; and 4) repeat sepsis admissions (only the first ICU stay was analyzed).

**Figure 1.**
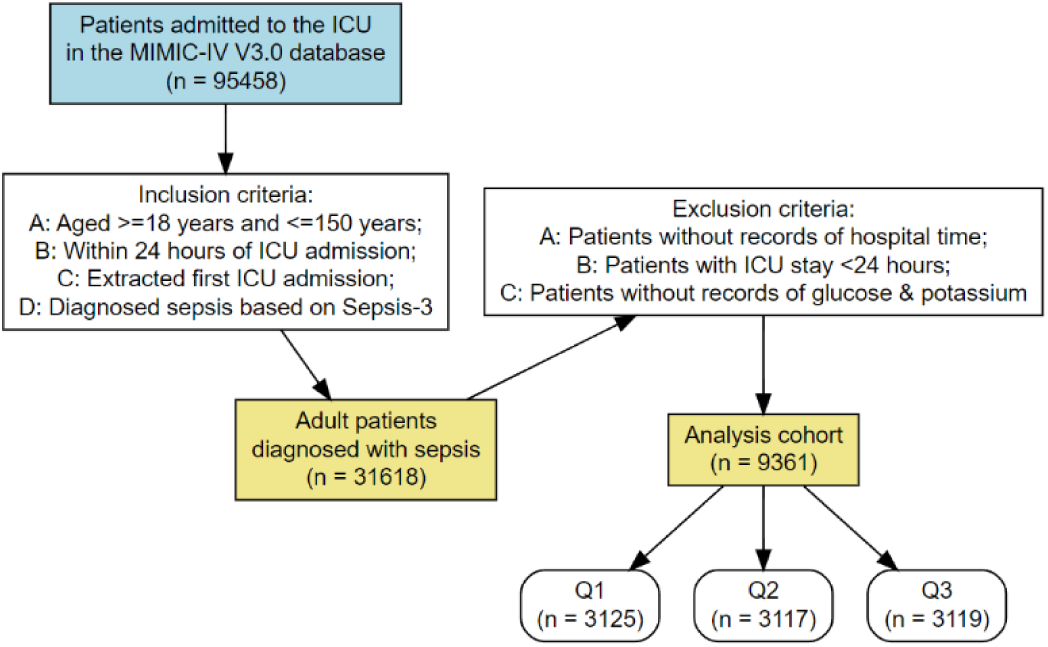
Flowchart of study population inclusion and exclusion.

### Data Extraction

Structured Query Language (SQL) via Navicat Premium v17.0 facilitated extraction of: Demographic variables: gender, age, height (meters), weight (kilograms), ethnicity; clinical severity scores: baseline Glasgow Coma Score, Sequential Organ Failure Score (SOFA), Simplified Acute Physiology Score (SAPS)-II, Acute Physiology Score (APS)-III, Oxford Acute Severity of Illness Score (OASIS), Systemic Inflammatory Response Syndrome Score ( SIRS); baseline vital signs: mean blood pressure (mmHg), mean heart rate (beats/min), systolic blood pressure (SBP, mmHg), respiratory rate (beats/min), diastolic blood pressure (mmHg), and pulse oximetry (SPO2, %); and laboratory variables: red blood cells (RBC, 10^9/L), hemoglobin (Hb, g/L), platelets (10^9/ L), white blood cells (WBC, 10^9/L), sodium (mmol/L), blood urea nitrogen (BUN, mg/dL), creatinine (mg/24 hours), triglycerides (TG, mg/dL), fasting blood glucose (FBG, mg/dL), potassium (mmol/L), anion gap (mmol/L), prothrombin time (sec), activated partial thromboplastin time (sec), international normalized ratio; treatments: mechanical ventilation (MT), vasopressors, oxygen; comorbidities: hypertension (HTN), diabetes mellitus (DM), heart failure (HF), cardiac arrhythmia, acute myocardial infarction (AMI), peripheral vascular disease, chronic obstructive pulmonary disease, respiratory failure, ventilator-associated pneumonia (VAP), chronic kidney disease, hyperlipidemia, malignancy, renal failure, sepsis, liver disease, and Charlson comorbidity index.

### Clinical outcomes

Primary outcomes included 90-day, 180-day and 1-year ACM. Secondary outcomes encompassed in-ICU mortality, in-hospital mortality, and 28-day ACM. All laboratory values were recorded as first measurements post-ICU admission. Variables with <20% missingness were addressed via multiple imputation.

### Statistical analyses

Normality of continuous variables was assessed using preliminary tests. For normally distributed data, Student’s t-test or one-way ANOVA was applied, with results expressed as mean ± standard deviation (SD). Non-normally distributed variables were analyzed using the Wilcoxon rank-sum test and reported as median (interquartile range, IQR). Categorical variables were compared using chi-square or Fisher’s exact tests, presented as absolute counts and proportions.

Kaplan-Meier (K-M) curves were generated to evaluate the cumulative incidence rates of primary and secondary outcomes across GPR quartiles. Univariate Cox regression was initially used to explore associations between GPR and short-/long-term all-cause mortality (ACM). Subsequently, multivariate Cox proportional hazards models were constructed, incorporating clinically relevant covariates or variables showing significance in univariate analysis. Covariate selection prioritized the availability of event data. Three hierarchical models were defined: Model 1 (unadjusted), Model 2 (adjusted for age, sex, and race), and Model 3 (further adjusted for hypertension (HTN), diabetes mellitus (DM), heart failure (HF), white blood cell count (WBC), red blood cell count (RBC), systolic blood pressure (SBP), and Sequential Organ Failure Assessment (SOFA) scores), with the lowest GPR quartile as the reference. Restricted cubic spline (RCS) analysis was used to examine GPR as a continuous variable to characterize its dose-response relationship with outcome risks. For nonlinear associations, recursive algorithms identified inflection points between GPR and ACM outcomes. Stratified analyses were conducted by age (<60 vs. ≥60 years), sex, race/ethnicity, HTN, and DM status. All analyses were performed using R 4.4.2 (The R Foundation), with statistical significance set at P < 0.05.

## Results

Table 1 presents the baseline demographic and clinical attributes stratified by GPR tertiles. Subjects were allocated into three tertiles based on their GPR values (Q1: ≤25.35; Q2: 25.35–34.69; Q3: ≥34.69). Subjects in the highest GPR tertile (Q3) exhibited higher incidences of chronic kidney disease (34.69% vs Q1: 30.73%), heart failure (42.42% vs Q1: 39.66%), and diabetes (58.03% vs Q1: 22.31%) compared to those in the lower tertiles (Q1 and Q2). Additionally, they had higher physiological markers, including heart rate (Q3: 88 vs Q1: 86 beats/min) and systolic blood pressure (Q3: 127 vs Q1: 124 mmHg). Oxygen partial pressure showed a non-linear pattern across tertiles (Q3: 102 mmHg vs Q2: 109 mmHg vs Q1: 105.5 mmHg). Laboratory indicators such as white blood cell count were elevated in the higher GPR tertiles (Q3: 10.00 vs Q1: 8.20 ×10⁹/L), while creatinine levels showed significant differences across tertiles (Q3 median: 1.00, IQR: 0.80-1.40 vs Q1 median: 1.00, IQR: 0.80-1.30, P<0.001). Clinical scores, including the Simplified Acute Physiology Score II (SAPS II) and the Oxford Acute Illness Severity Score (OASIS), were higher in the higher GPR tertiles, indicating greater disease severity. Furthermore, therapeutic measures showed increased use of invasive ventilation (Q3: 56.84% vs Q1: 50.97%), with subjects in the highest GPR tertile requiring longer durations of ventilation (Q3: 20.55 vs Q1: 18.48 hours). Mortality data showed that subjects in the highest GPR tertile had lower 30-day mortality (Q3: 6.83% vs Q1: 9.38%, P<0.001), reflecting better outcomes despite their higher baseline disease burden. Other notable differences included higher prevalence of hypertension in Q3 (61.08% vs Q1: 55.19%), lower malignancy rates in Q3 (18.05% vs Q1: 24.65%), and lower sodium levels in Q3 (138 vs 139 mmol/L). Race distribution showed significant variation (P<0.001), with a higher representation of other ethnic groups in Q3 (21.83% vs Q1: 16.68%).

**Table1.**
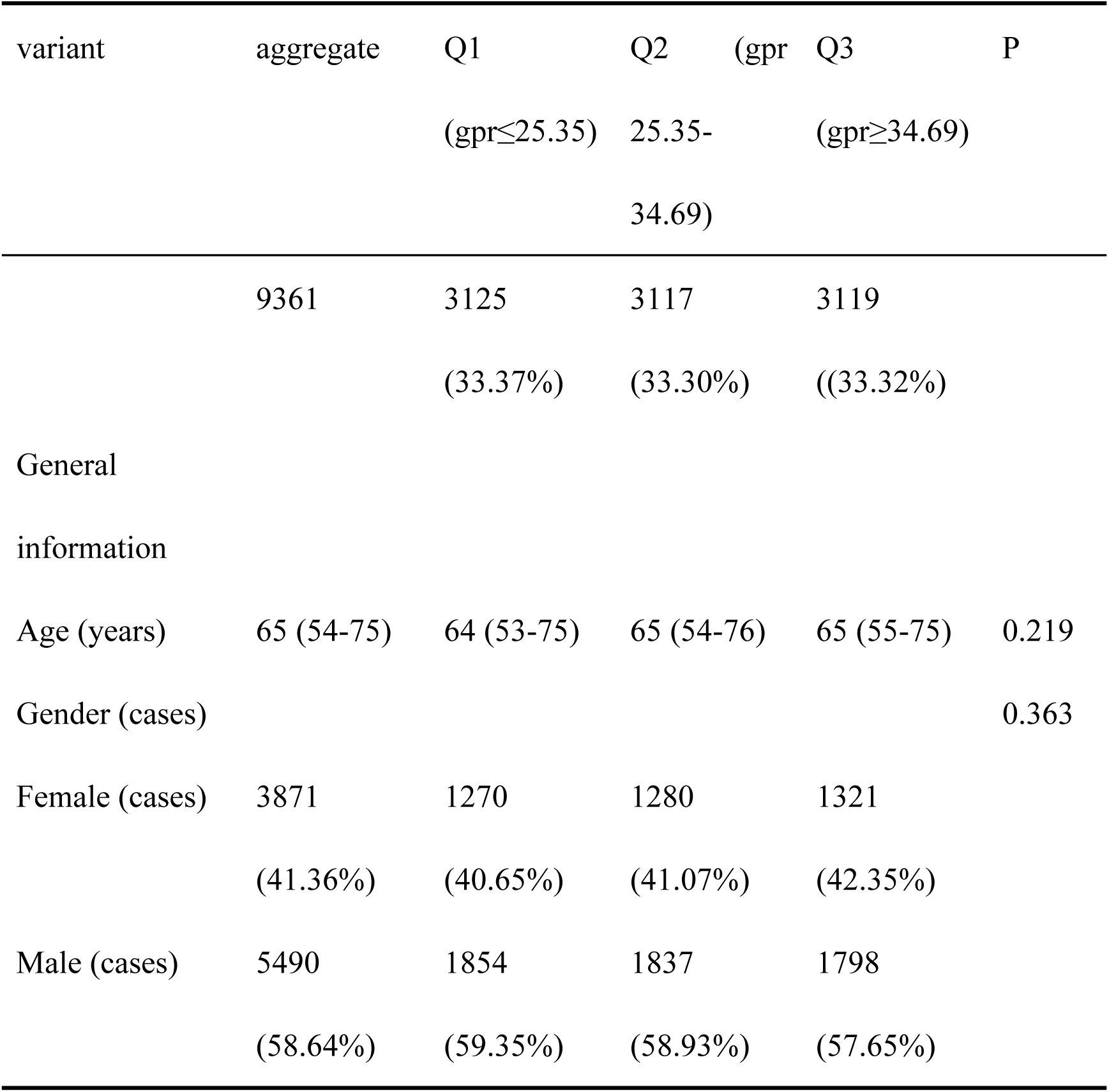
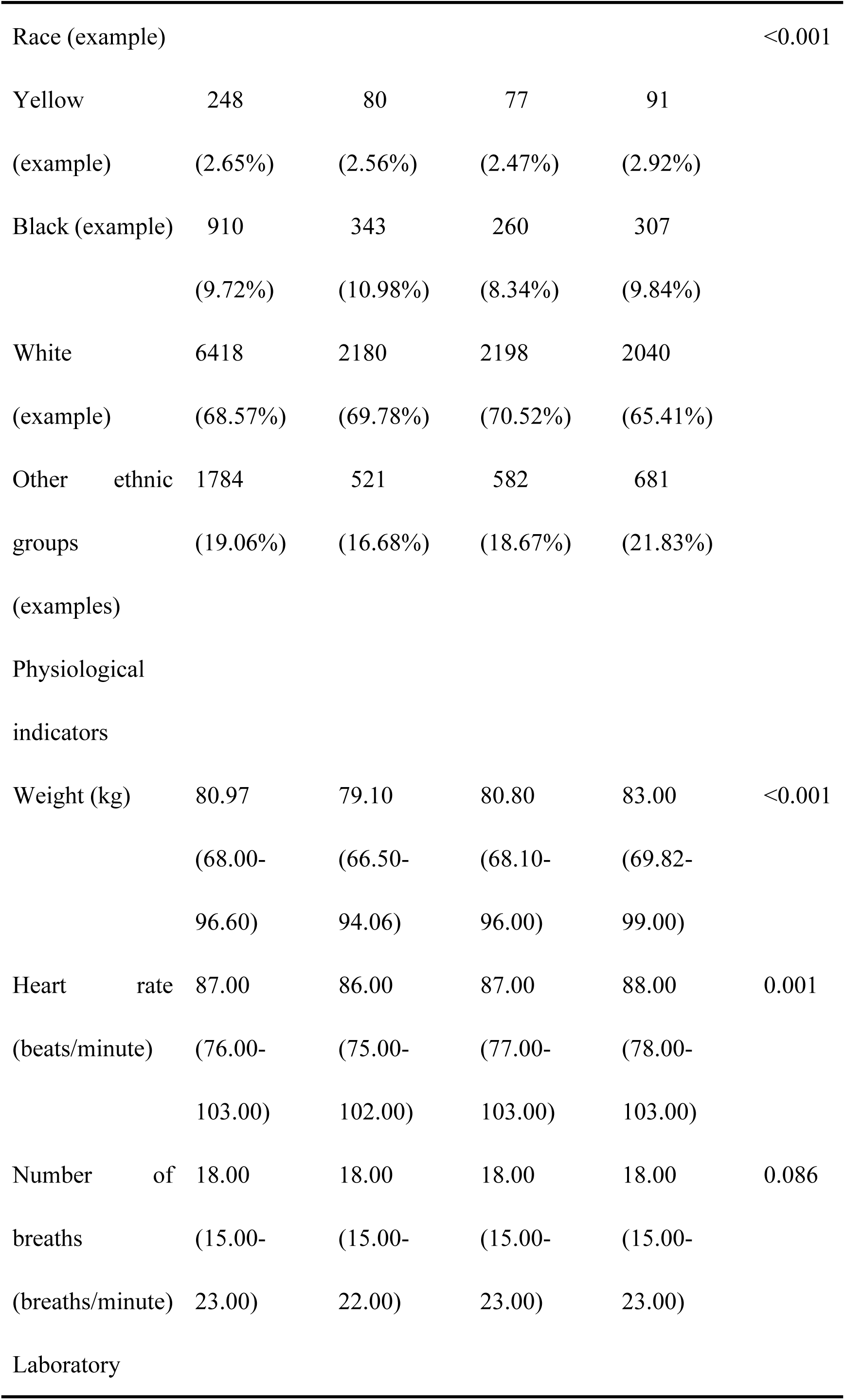
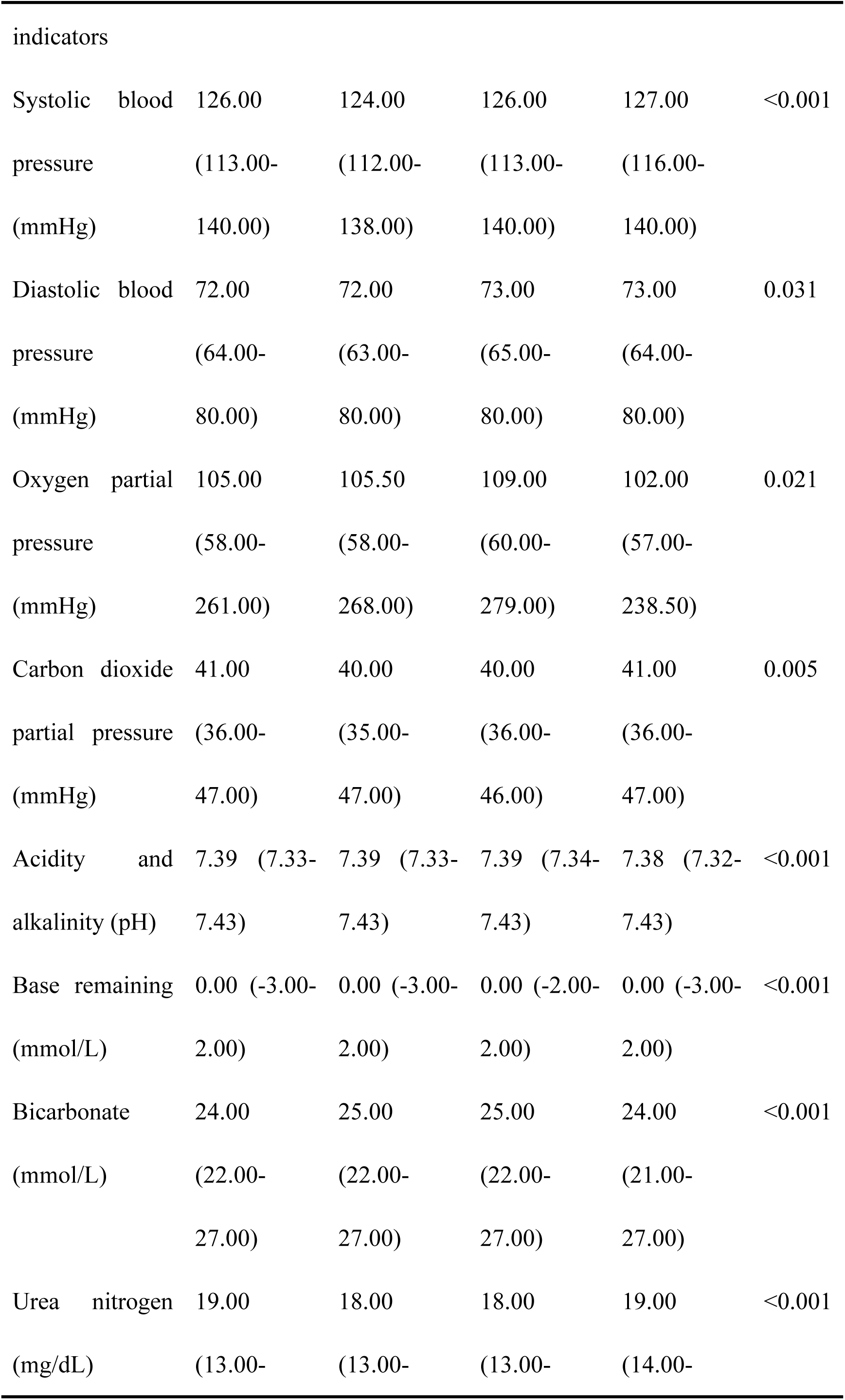
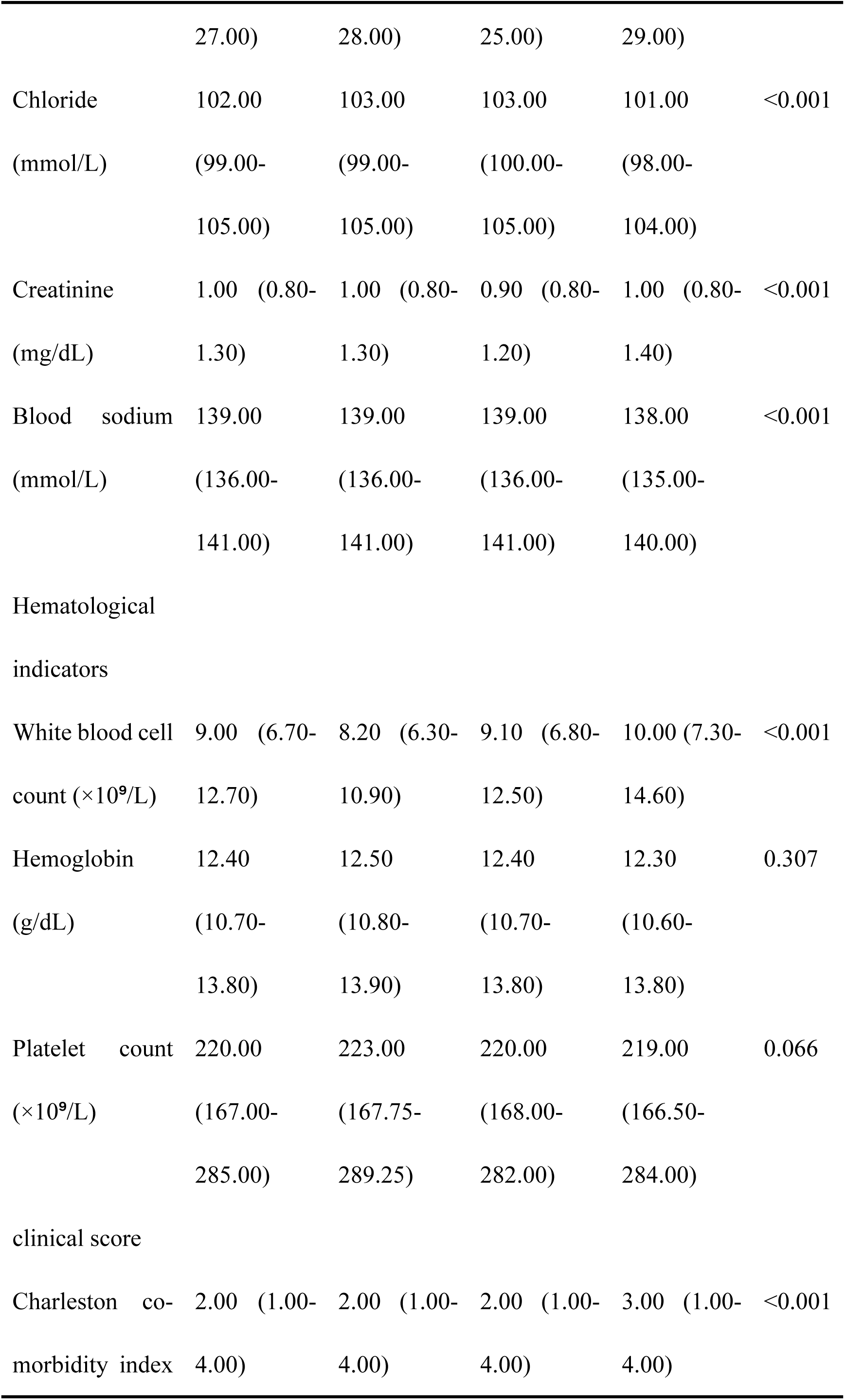
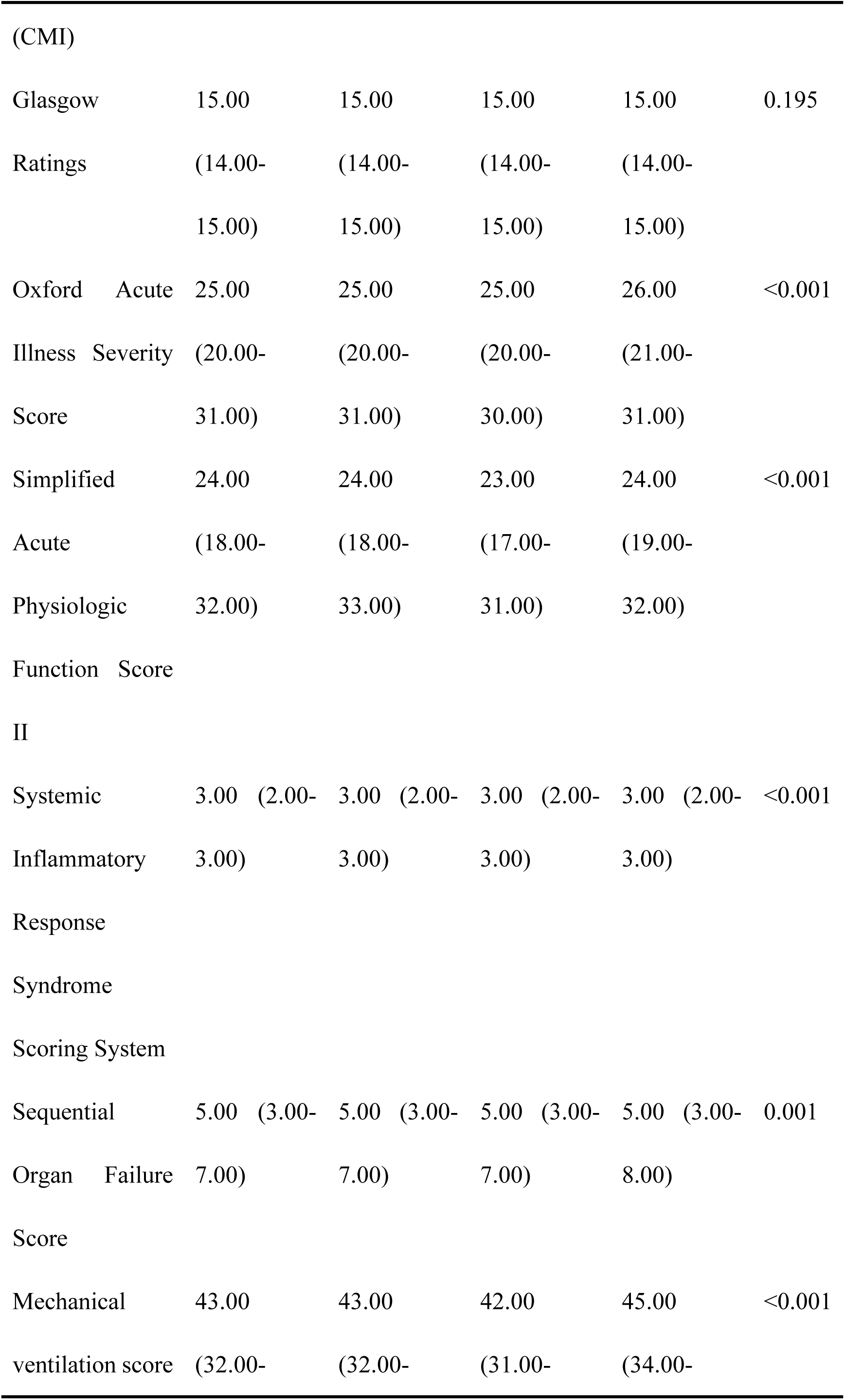
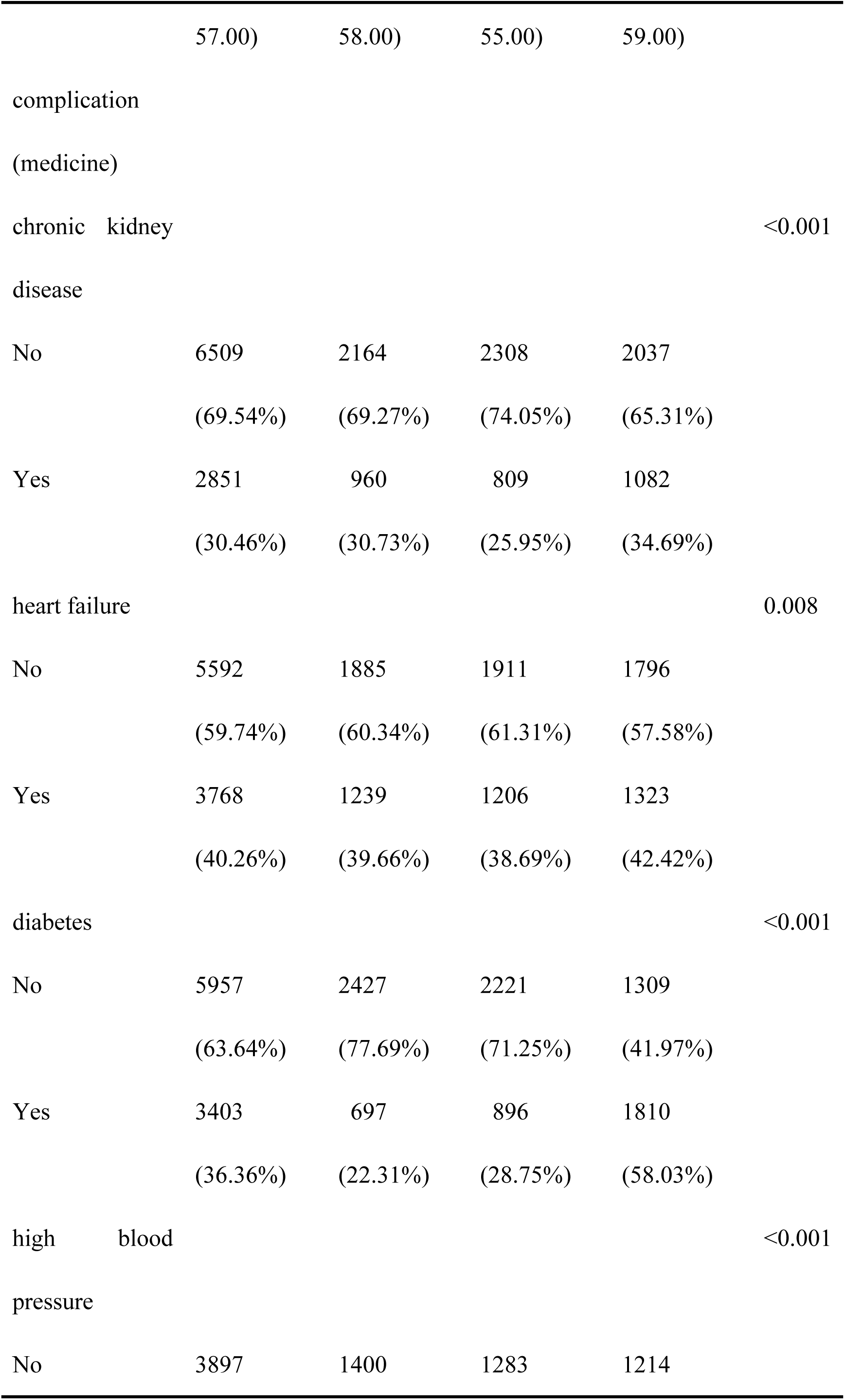
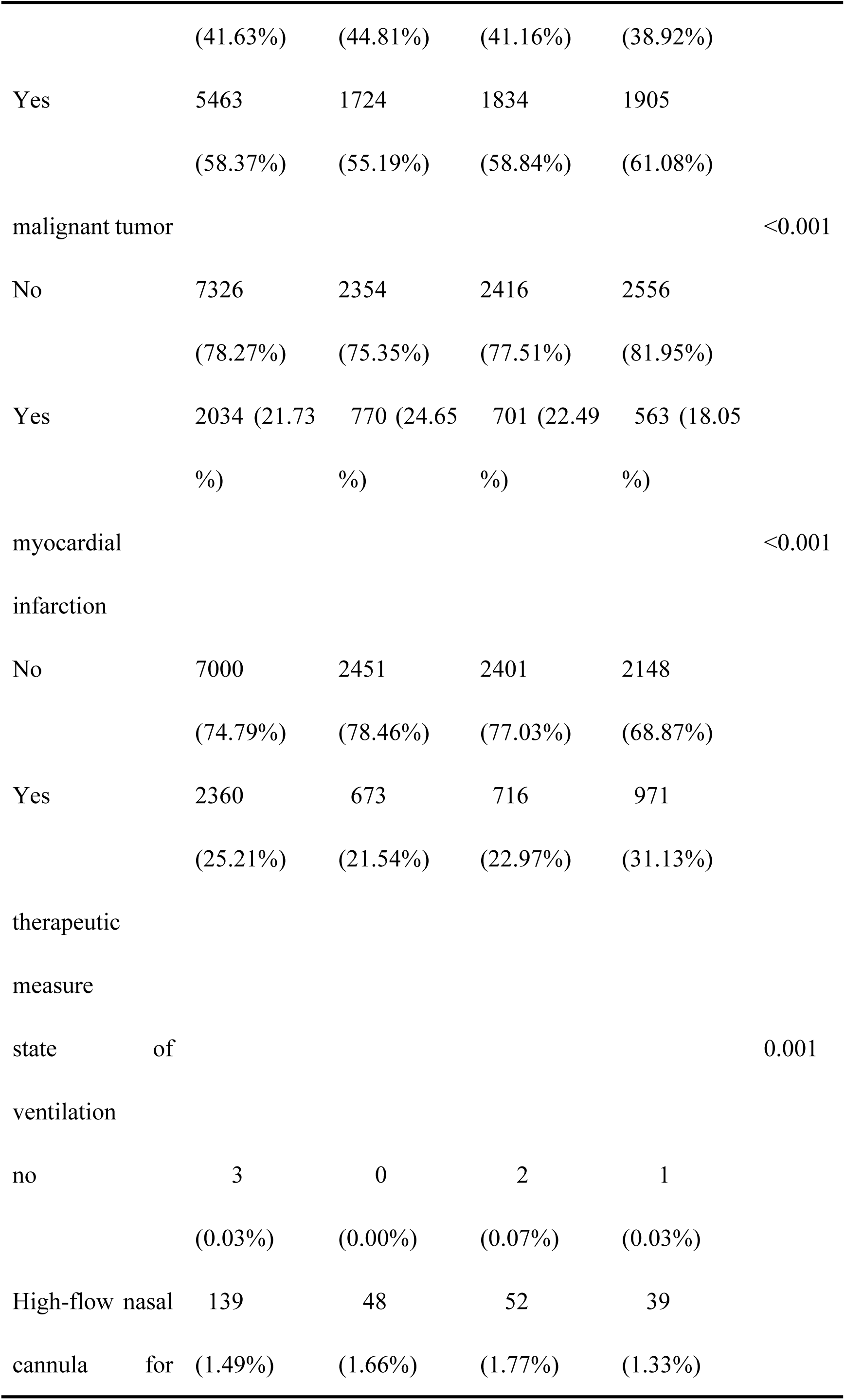
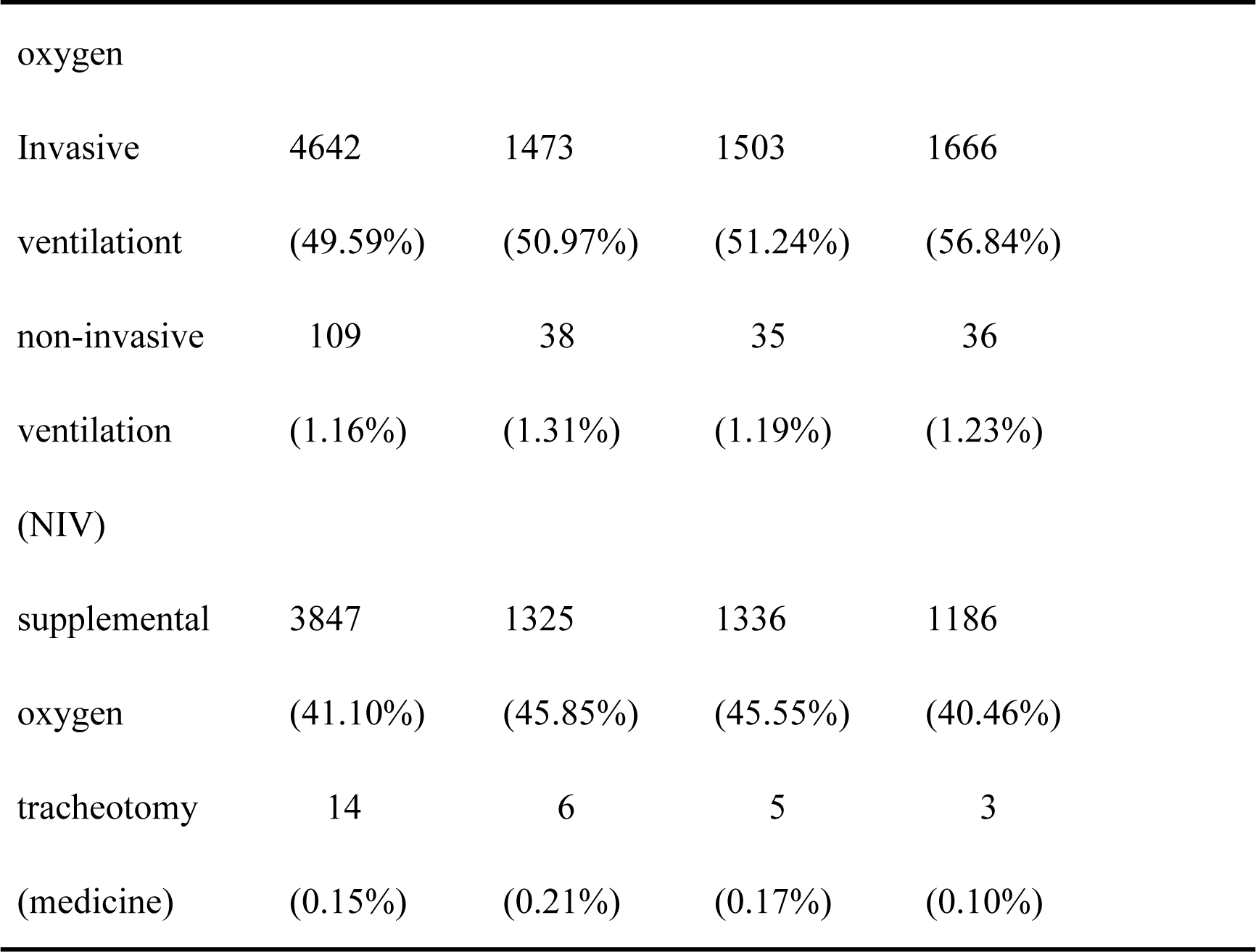
Baseline Characteristics of the Study Population.

### Survival Outcomes in Sepsis Patients Stratified by GPR Levels

To investigate the relationship between GPR levels and clinical outcomes in sepsis patients, we performed a Kaplan-Meier survival analysis, as shown in Figure 2. The results indicate that patients with lower GPR levels had a significantly lower probability of survival, whereas those with higher GPR levels demonstrated a higher probability of survival. The p-values for the log-rank test at 28 days, 90 days, 180 days, and 1 year were 0.00043, < 0.0001, 0.00019, and 0.0004, respectively, confirming a significant difference in survival across different GPR levels.

**Figure1.**
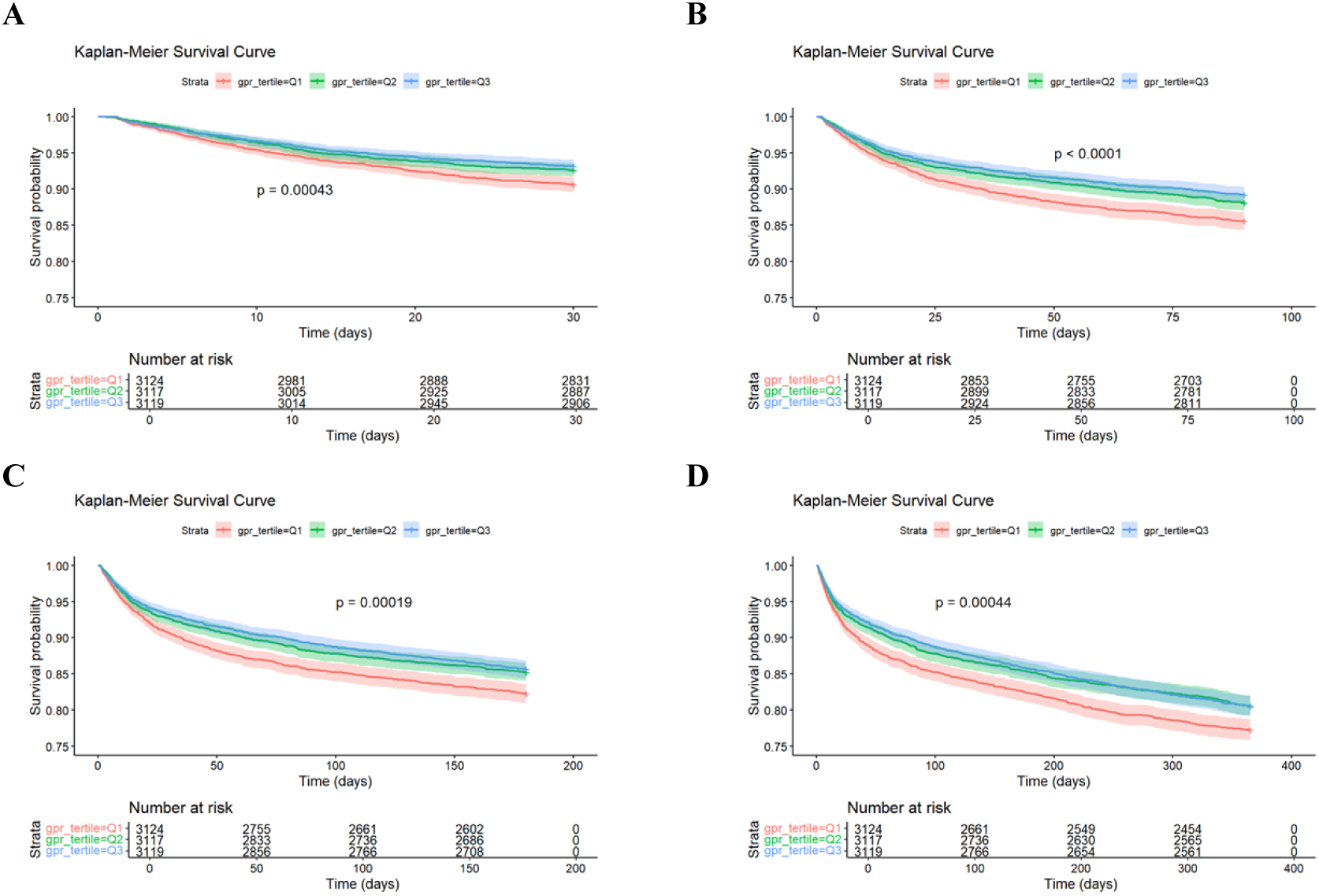
Kaplan-Meier curves for patients with sepsis. Note: A, B, C, and D are survival rates of septic patients at 28 days, 90 days, 180 days, and 1 year, respectively

### GPR correlates with clinical outcomes in patients with sepsis

We constructed three Cox regression models to assess the association between GPR levels and all-cause mortality (ACM) in sepsis patients. After fully adjusting for covariates (age, gender, ethnicity, comorbidities, and clinical indicators), higher GPR levels were consistently associated with a reduced mortality risk. In the fully adjusted model (Model 3), the highest GPR tertile (Q3) showed significantly lower mortality risk at 28 days (HR=0.712, 95% CI: 0.591-0.857, p<0.001), 90 days (HR=0.707, 95% CI: 0.610-0.821, p<0.001), 180 days (HR=0.752, 95% CI: 0.659-0.857, p<0.001), and 1 year (HR=0.788, 95% CI: 0.703-0.883, p<0.001). Trend tests were significant (p<0.0001), indicating a robust inverse relationship between GPR levels and mortality risk across all time points (Table 2).

**Table2.**
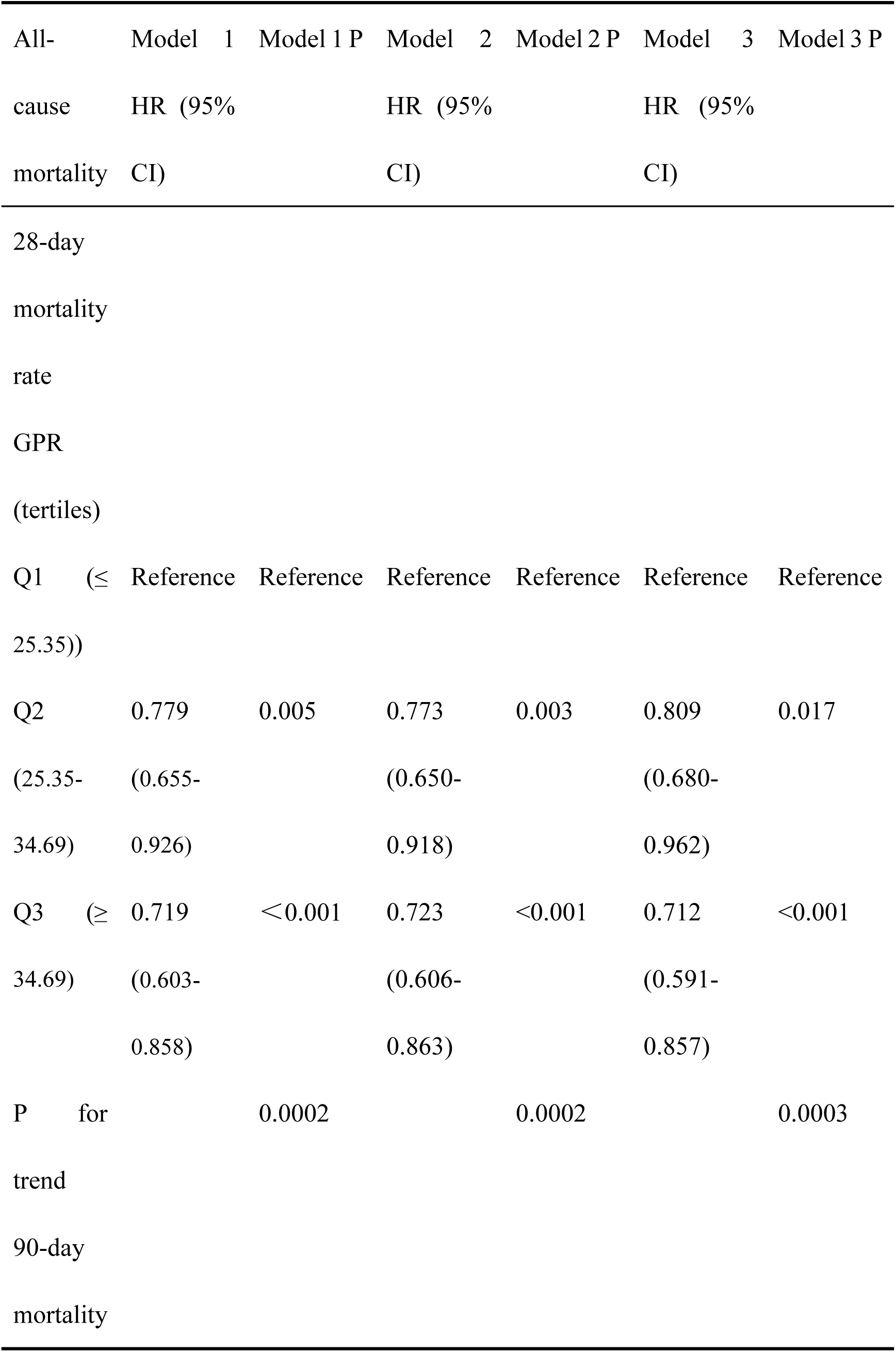
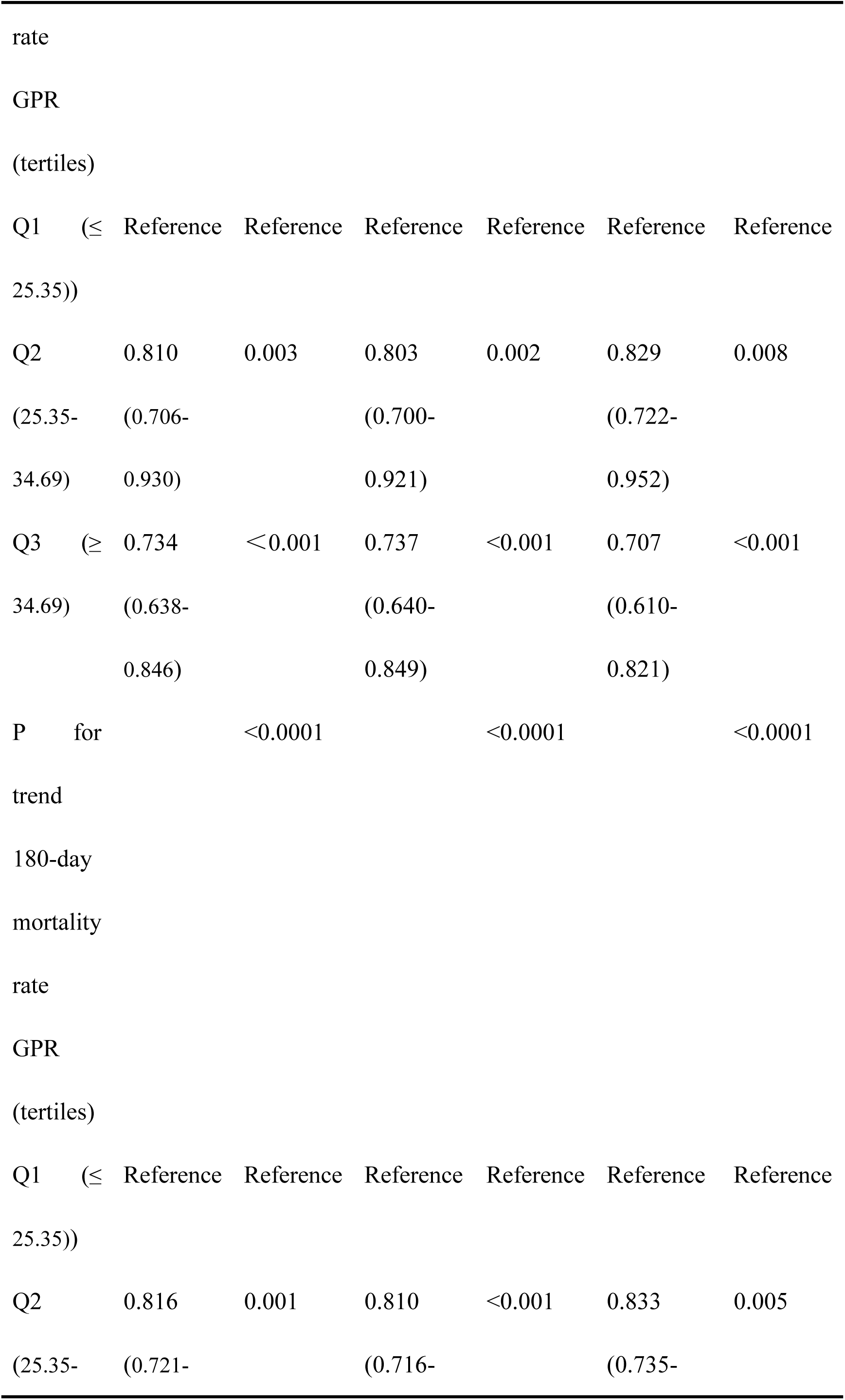
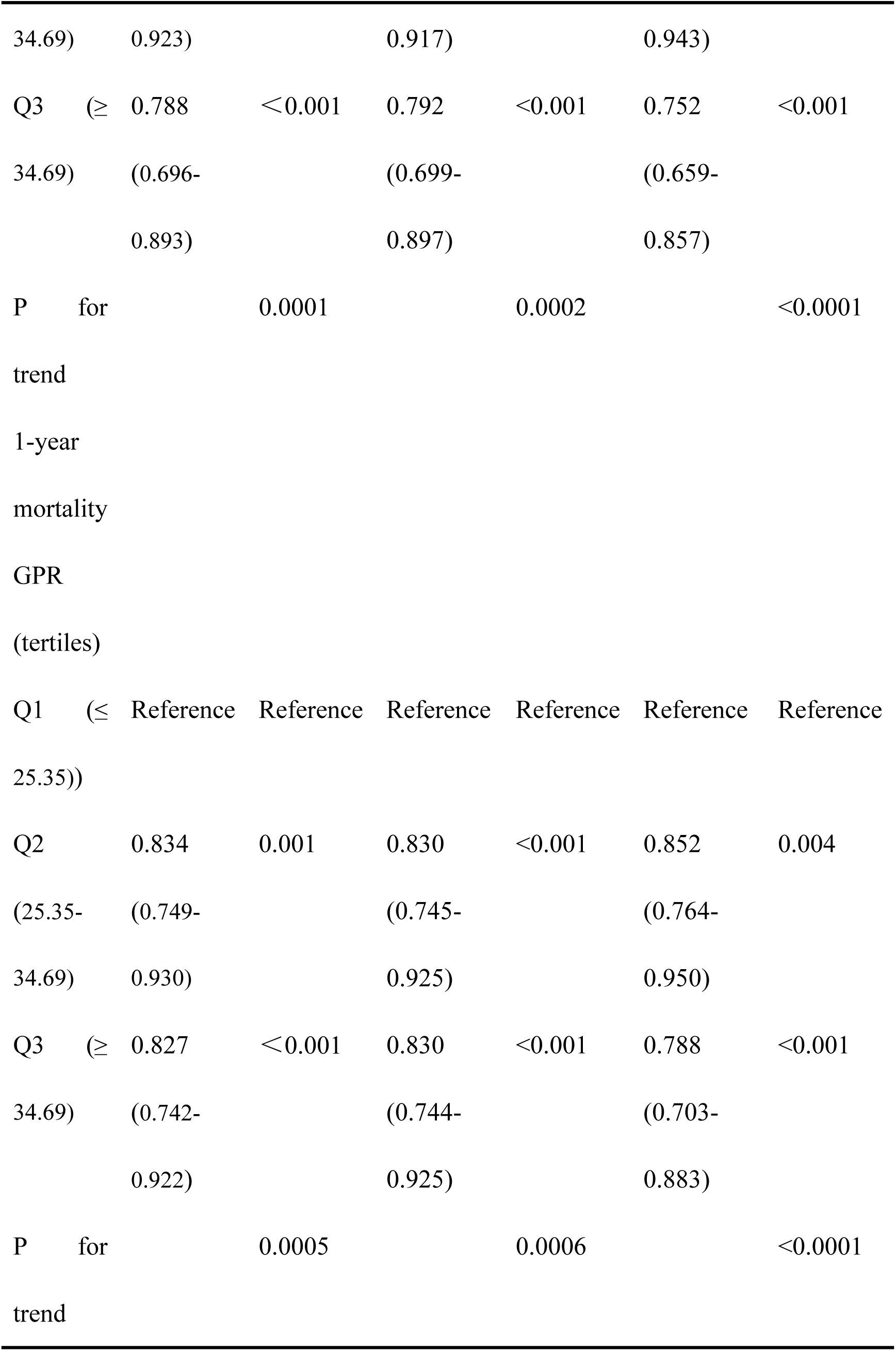
Multivariate Cox proportional risk models for long-term all-cause mortality.

### Nonlinear Relationship Between GPR and Survival in Sepsis Patients

Restricted cubic spline analysis revealed a significant L-shaped association between GPR and all-cause mortality (ACM) in sepsis patients (Figure 3). GPR exhibited a threshold effect, with mortality risk stabilizing above a specific threshold. This nonlinear relationship was significant at 28 days (p=0.0108), 90 days (p=0.00417), 180 days (p=0.0012), and 1 year (p=0.000183). Notably, when GPR exceeded 34.69 (Q3), 1-year mortality risk significantly decreased (HR=0.788, 95% CI: 0.703-0.883). These findings suggest a plateau in GPR’s protective effect, emphasizing the importance of identifying critical thresholds for clinical decision-making in both acute and long-term sepsis management.

**Fig. 3.**
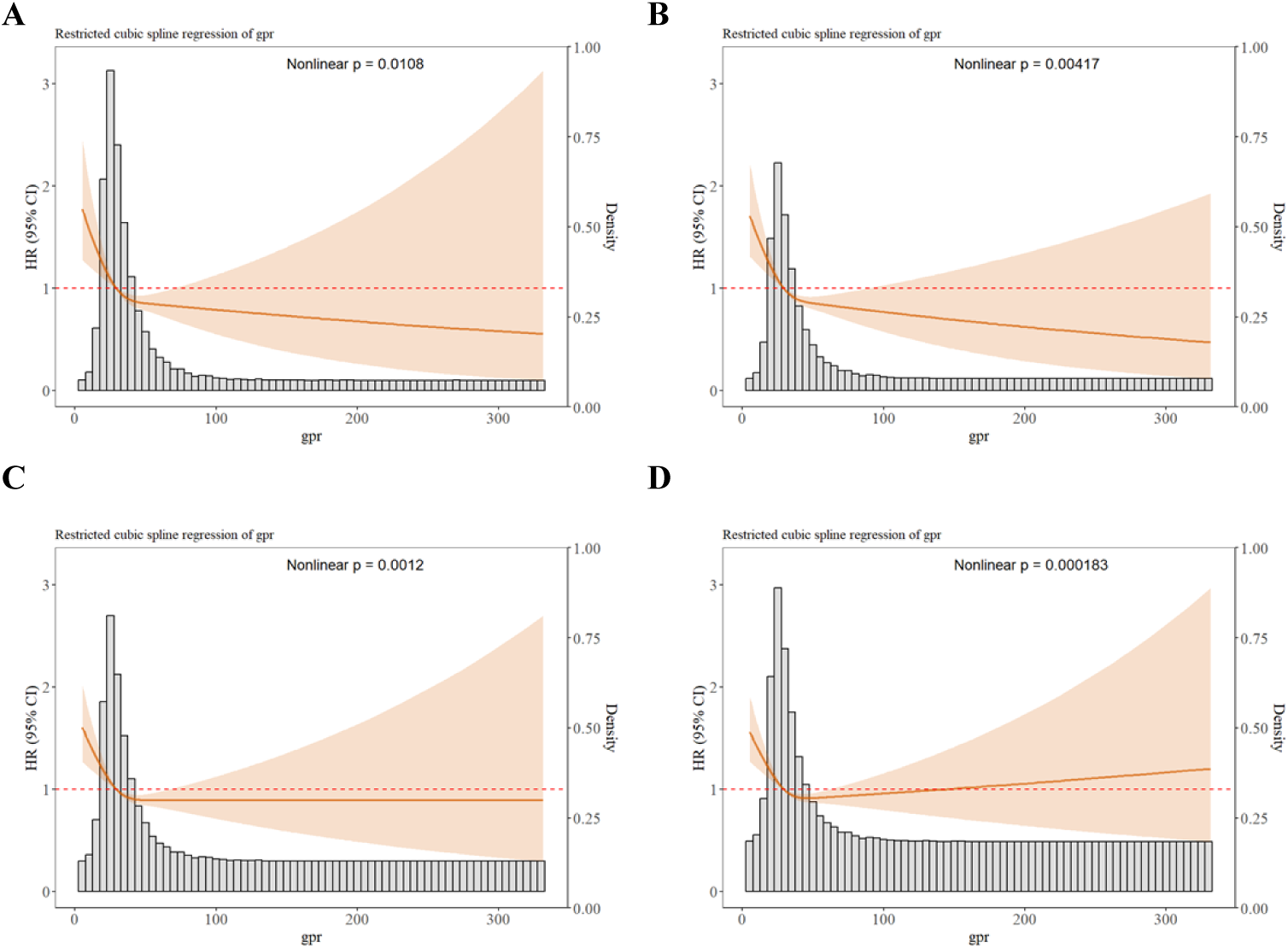
Restricted cubic spline analysis at different times. Note: A, B, C, and D indicate the relationship between GPR and ACM in septic patients at 28 days, 90 days, 180 days, and 1 year, respectively.

### Subgroup analysis

The prognostic value of GPR was further evaluated across multiple subgroups stratified by age, gender, race, comorbidities, and mechanical ventilation status (Fig. 4). GPR was significantly associated with a reduced risk of 28-day mortality in patients aged ≥60 years (HR: 0.990; 95% CI: 0.984–0.996, P<0.001), females (HR: 0.985; 95% CI: 0.977–0.993, P<0.001), and non-hypertensive patients (HR: 0.986; 95% CI: 0.978–0.994, P=0.005). Similarly, for 90-day mortality (Table 3), a significant protective effect was observed in older patients (HR: 0.972; 95% CI: 0.956–0.988, P<0.001) and females (HR: 0.983; 95% CI: 0.976–0.990, P<0.001). In racial subgroups, GPR showed a significant protective effect in Asians (28-day HR: 0.903; 95% CI: 0.835–0.976, P=0.033) and whites (90-day HR: 0.976; 95% CI: 0.958–0.995, P=0.001). Among patients receiving mechanical ventilation, GPR was associated with reduced 28-day mortality (HR: 0.993; 95% CI: 0.986–0.999, P<0.001). These findings highlight the consistent protective role of GPR across diverse patient subgroups.

**Figure 4.**
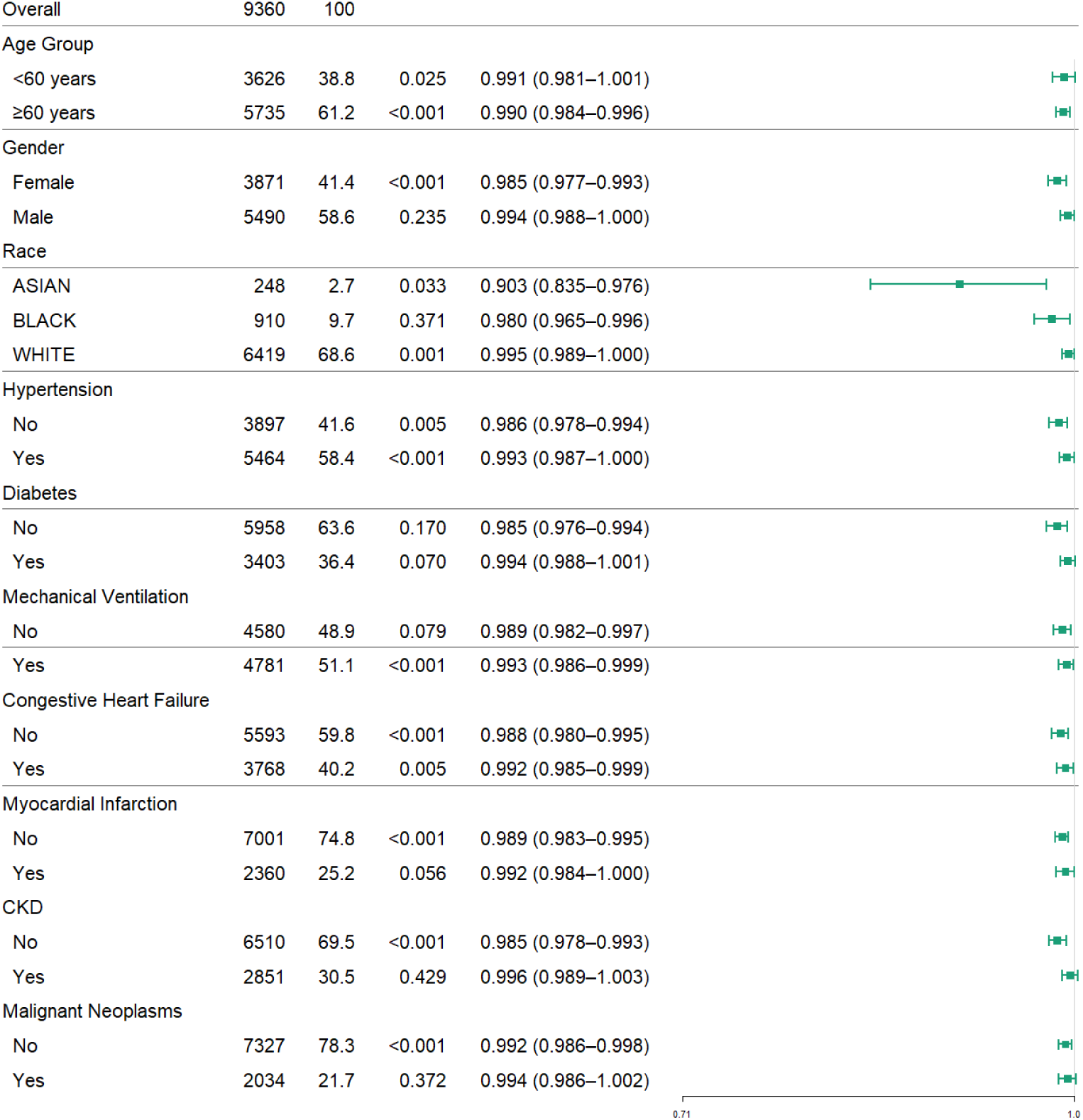
Subgroup analysis of 28-day ACM in patients with GPR and sepsis.

**Table3.**
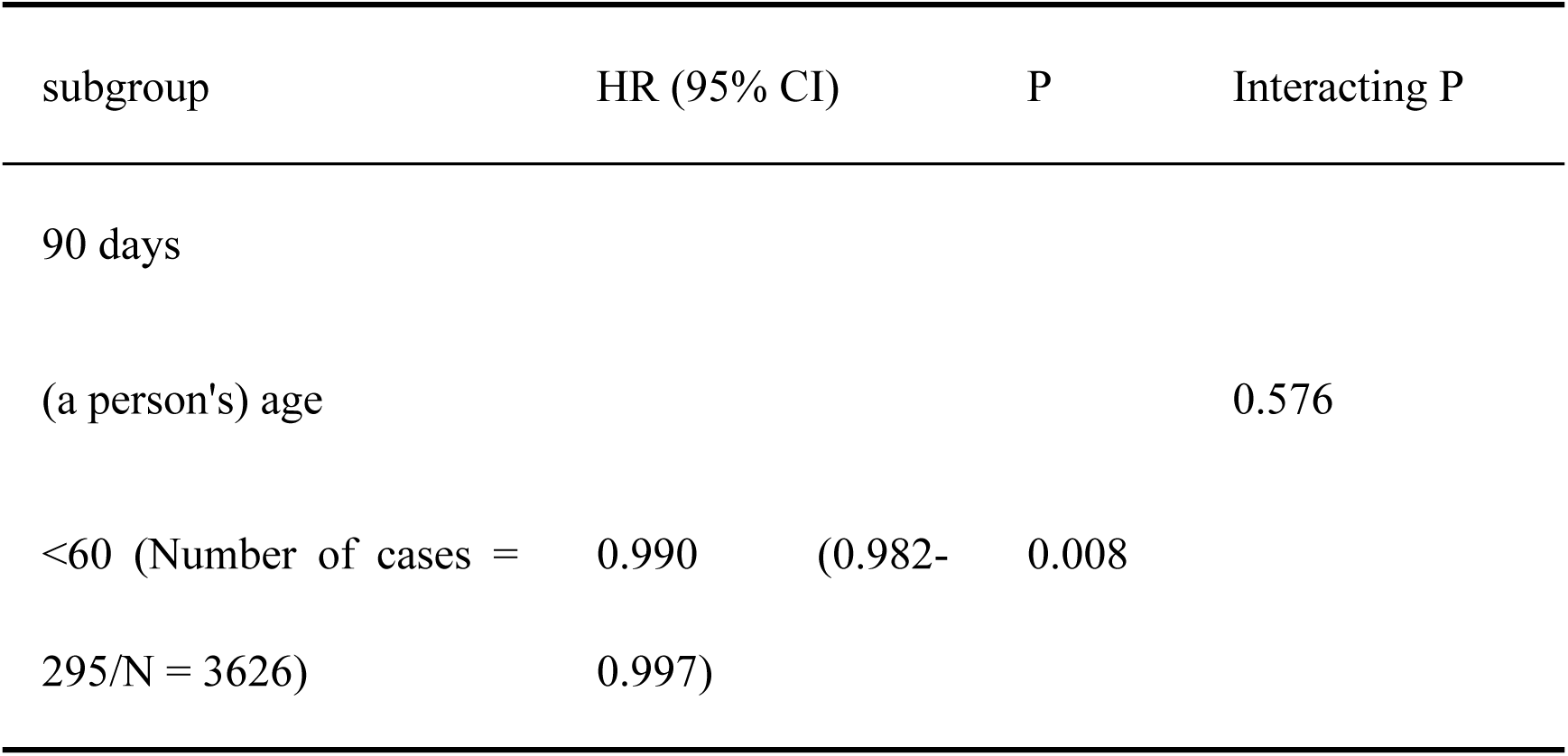
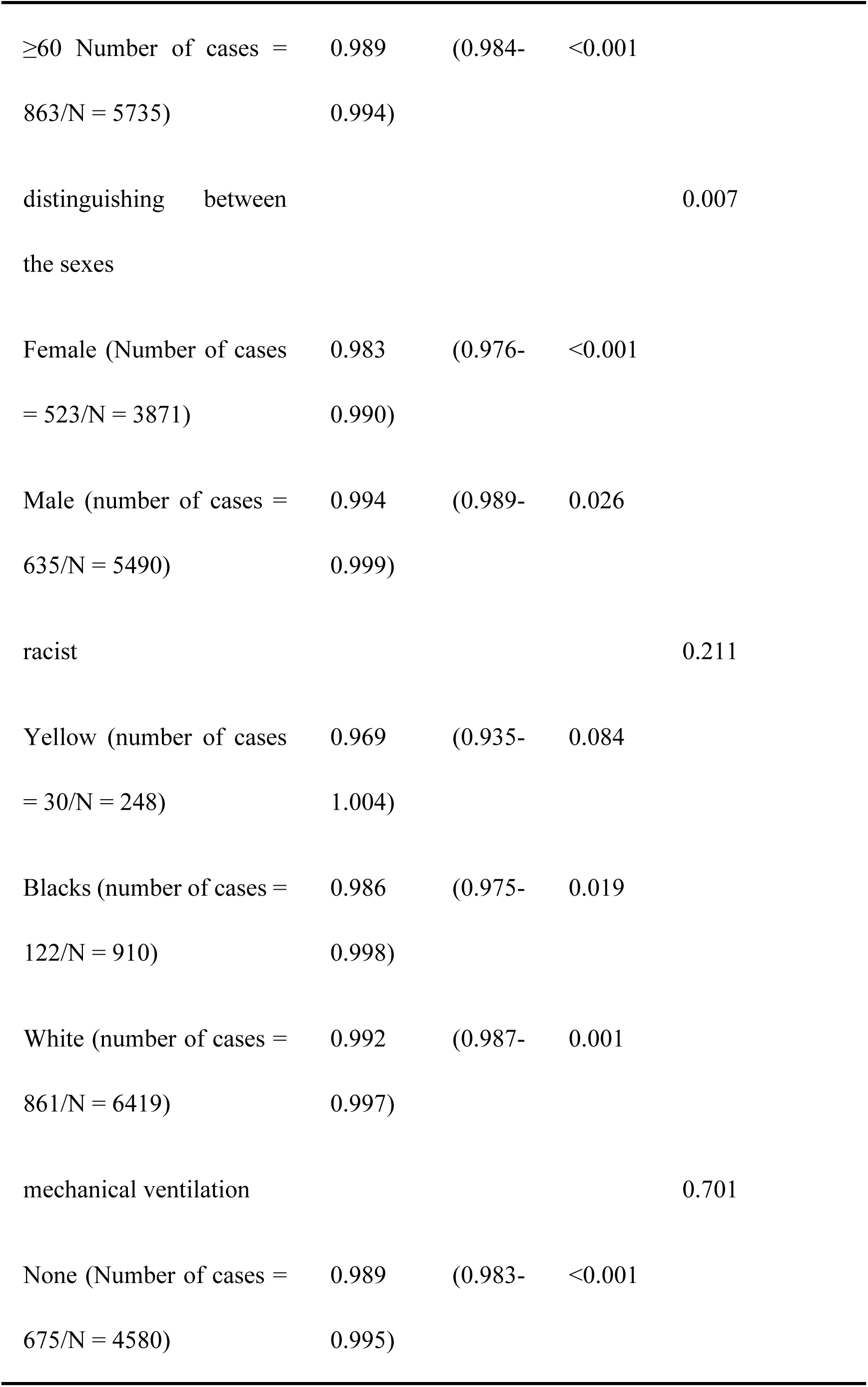
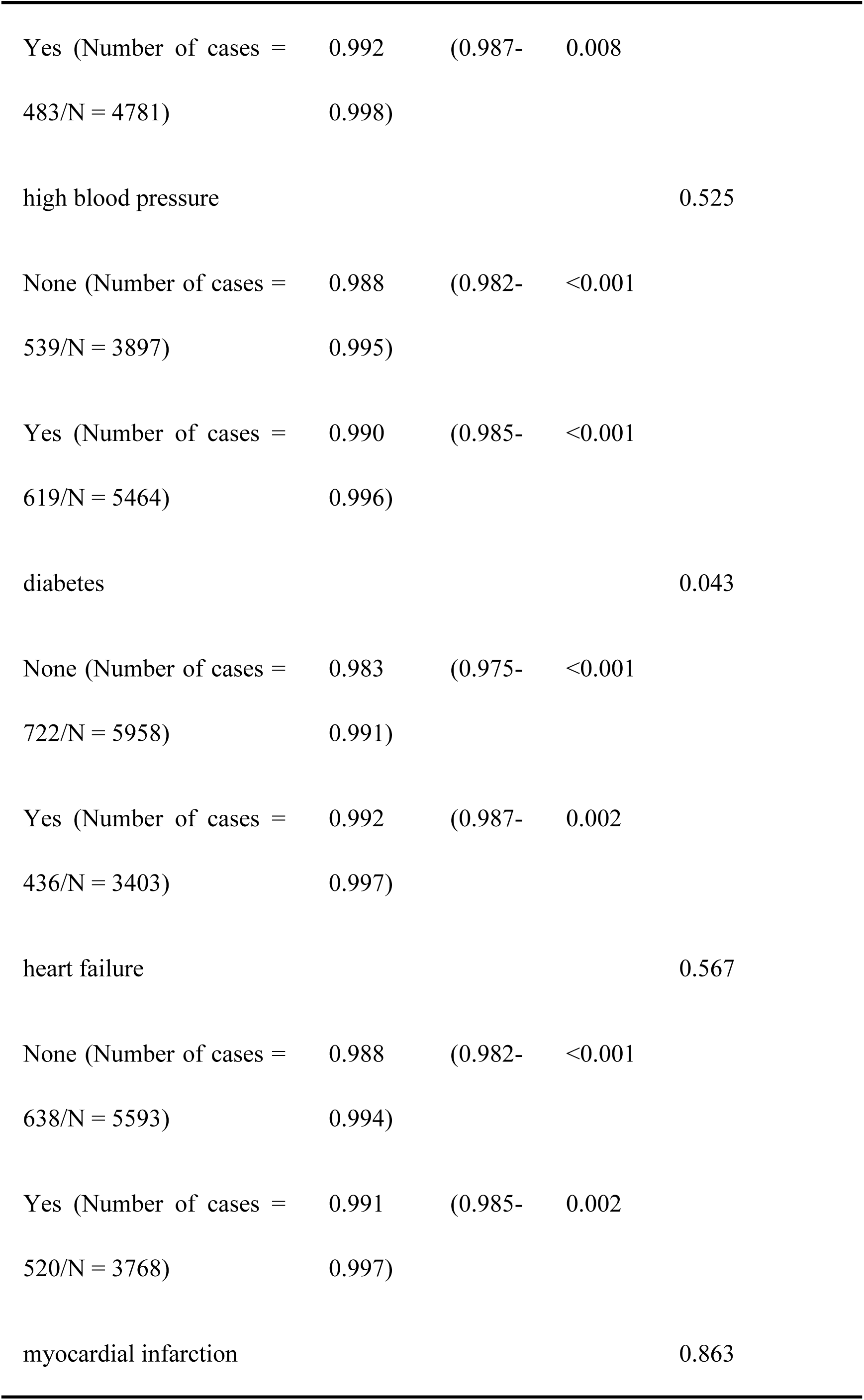
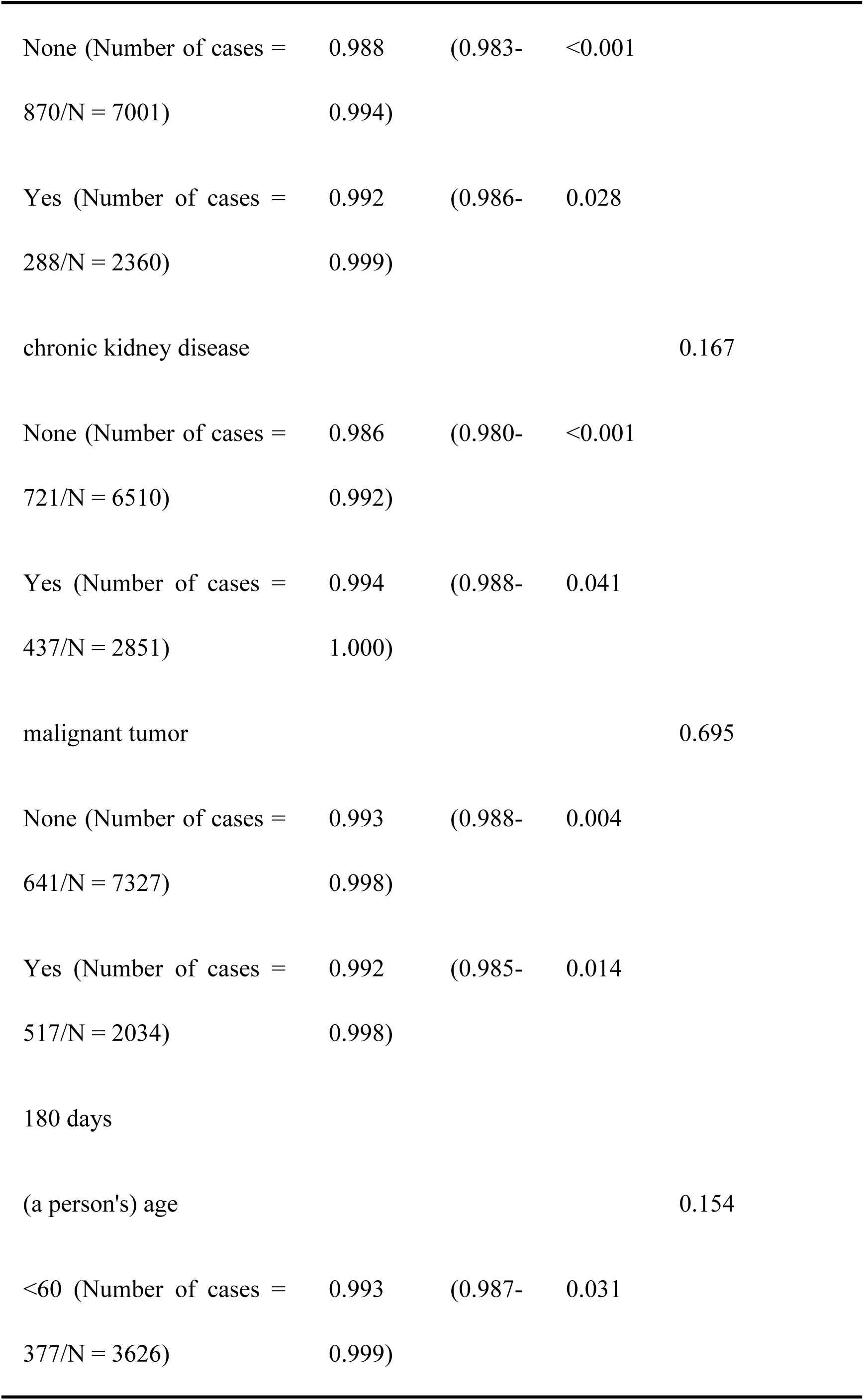
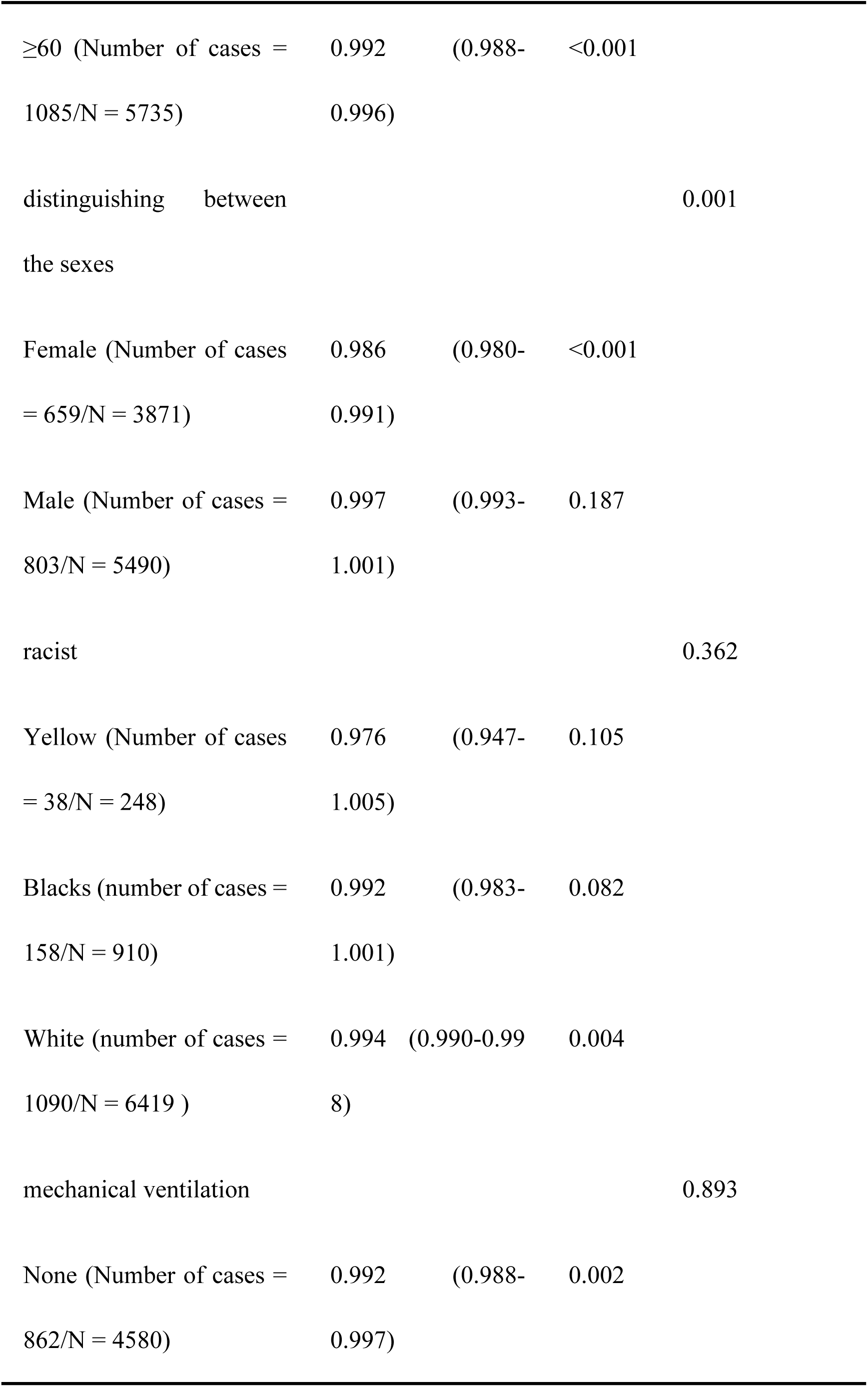
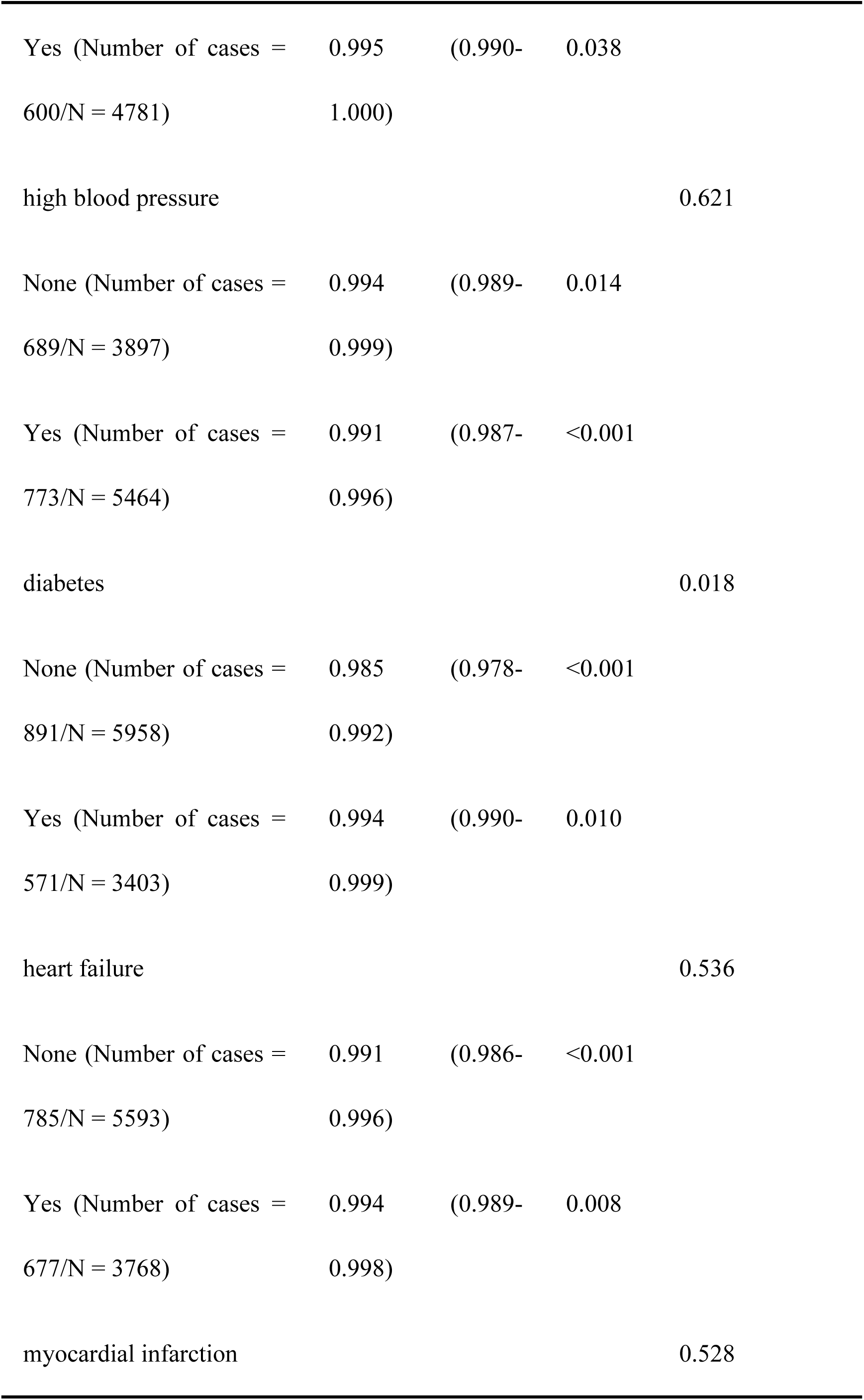
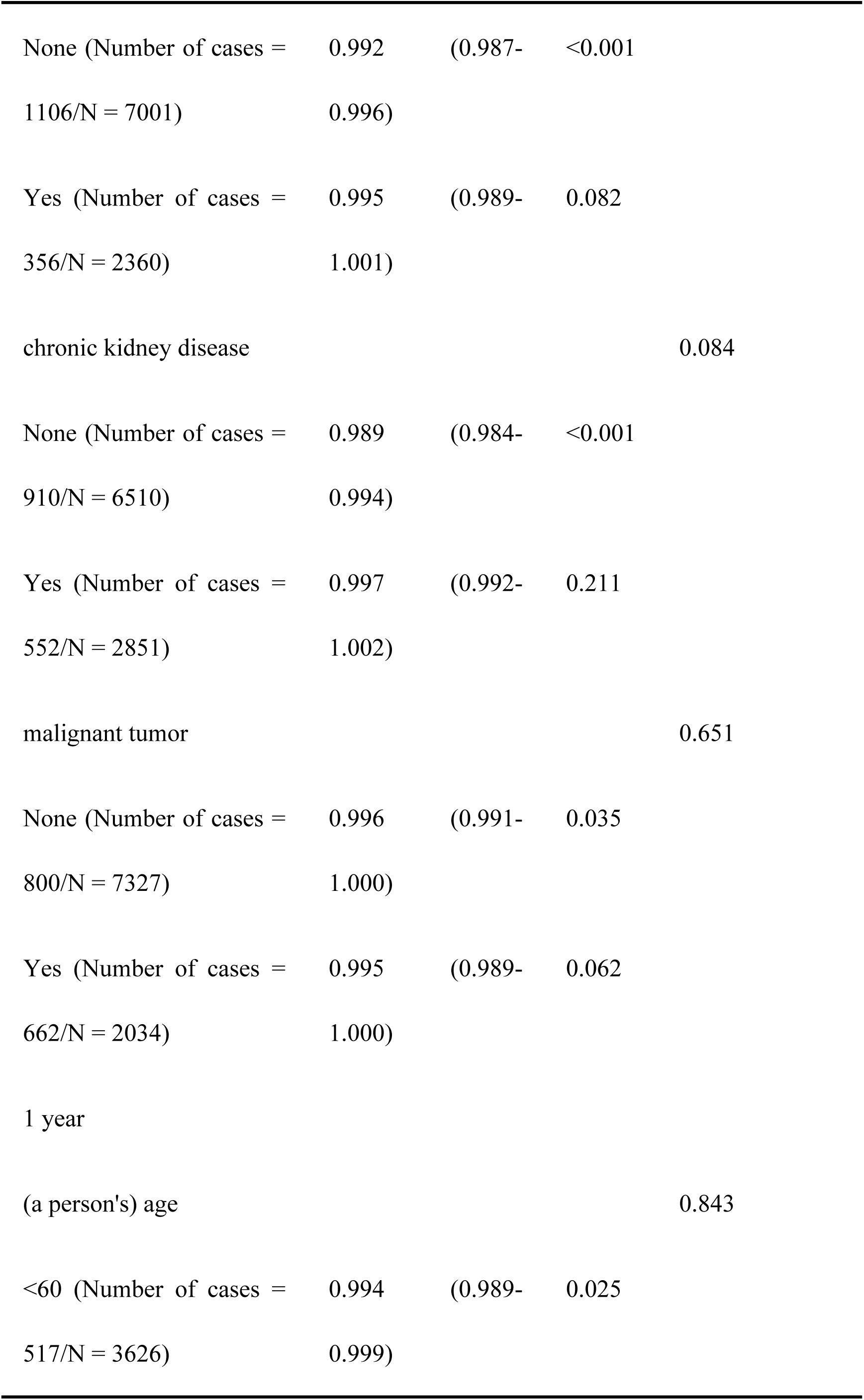
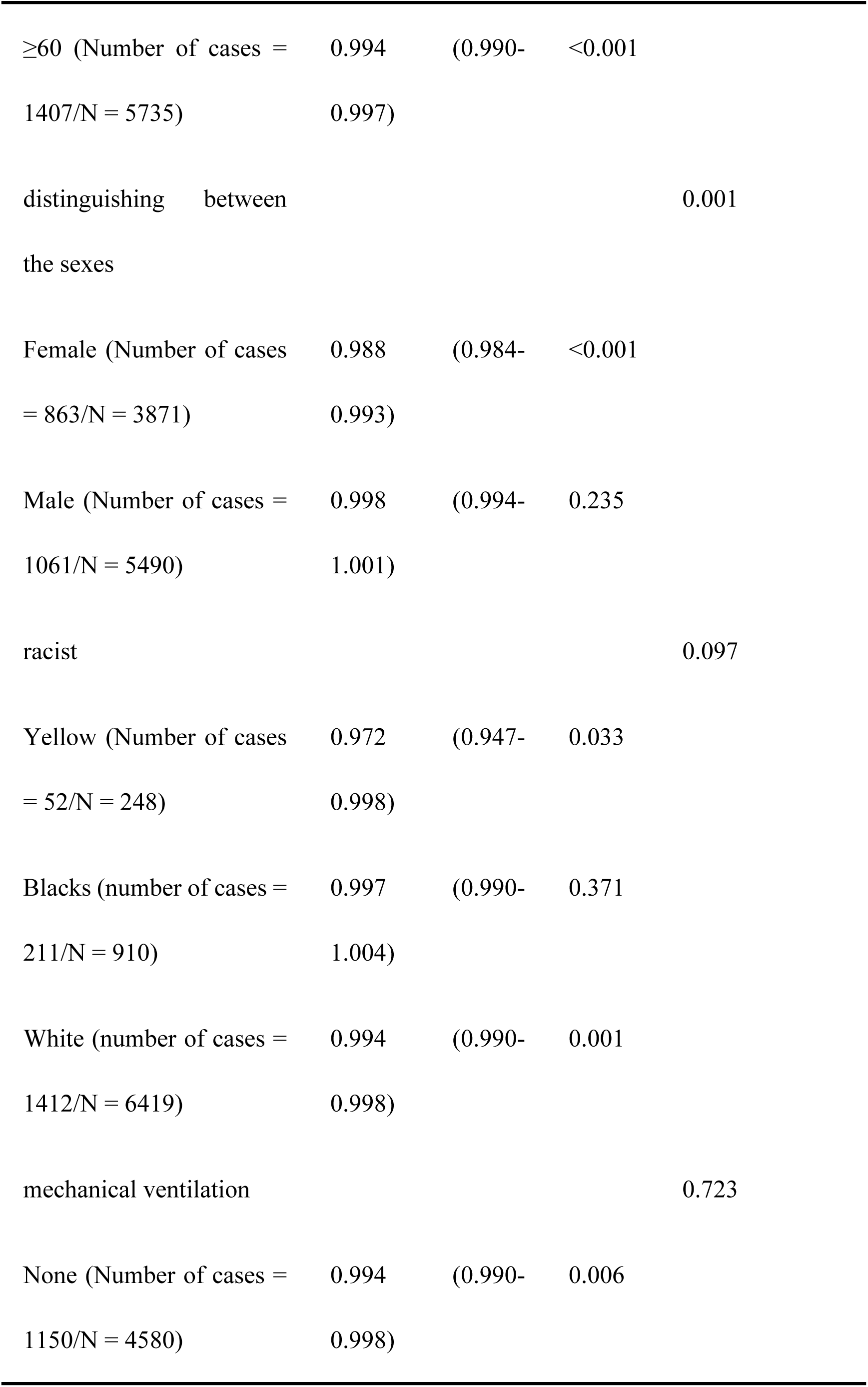
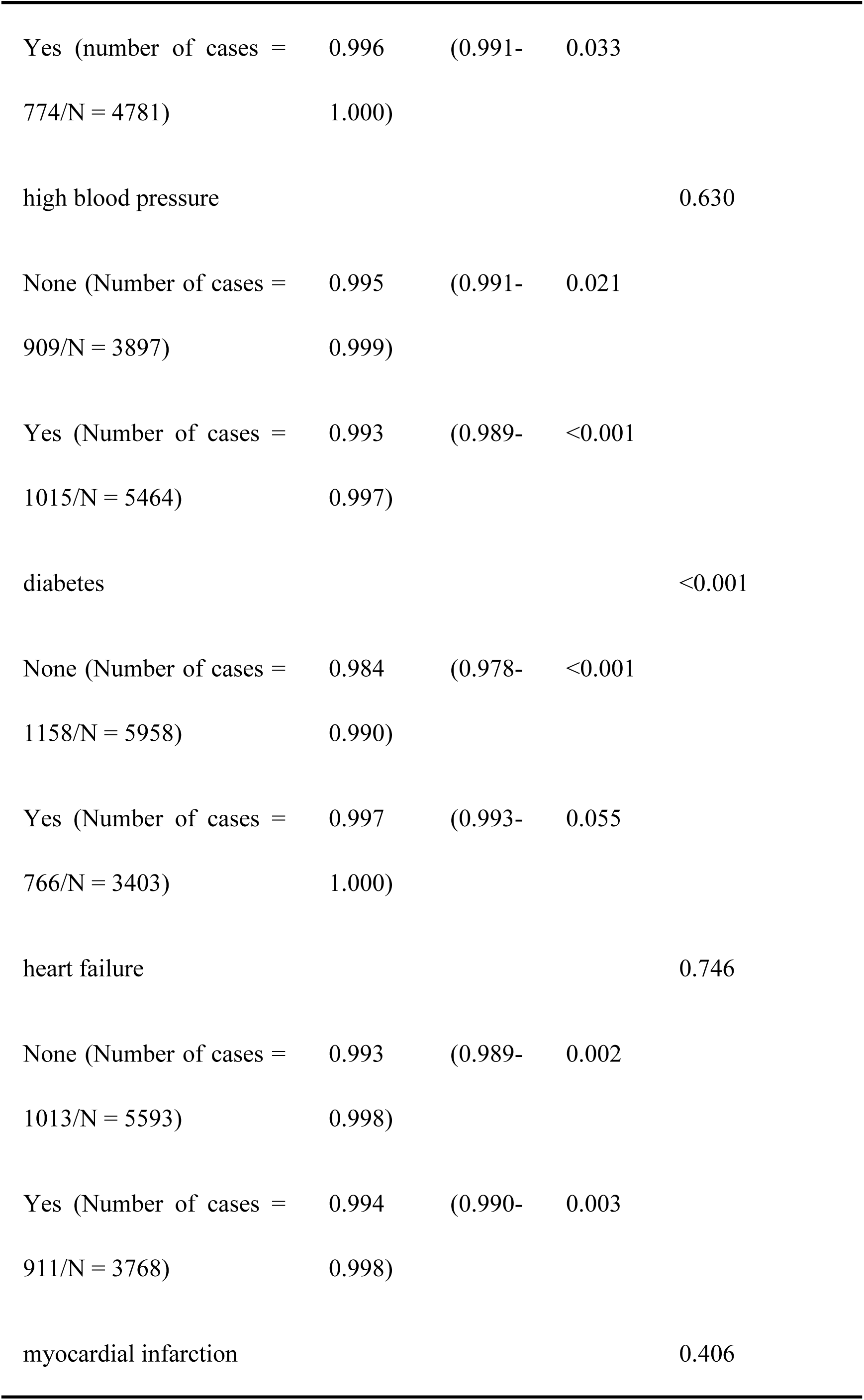
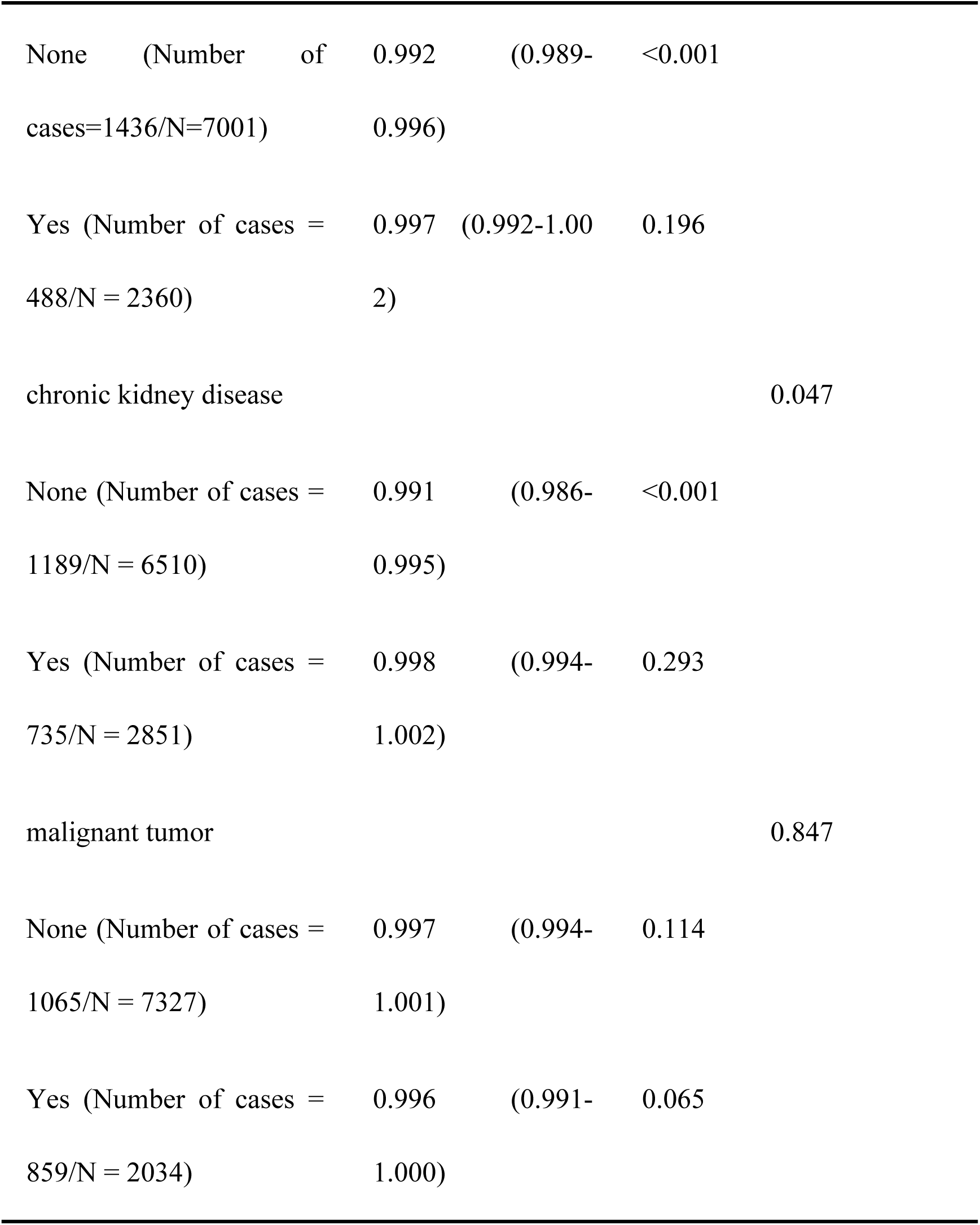
Subgroup analysis of GPR and long-term ACM.

### GPR Outperforms Glucose and Potassium in Predicting Sepsis Mortality

To evaluate the predictive ability of GPR for sepsis mortality, we compared the ROC curves of GPR, blood glucose, and blood potassium. As shown in Fig. 5, GPR demonstrated superior predictive performance, with AUCs of 0.677 (28 days), 0.645 (90 days), 0.626 (180 days), and 0.613 (1 year). In contrast, blood glucose (AUCs: 0.535–0.519) and potassium (AUCs: 0.516–0.526) showed significantly lower predictive accuracy across all time points. These results highlight that GPR is a more reliable predictor of mortality in sepsis patients compared to glucose or potassium alone.

**Fig. 5.**
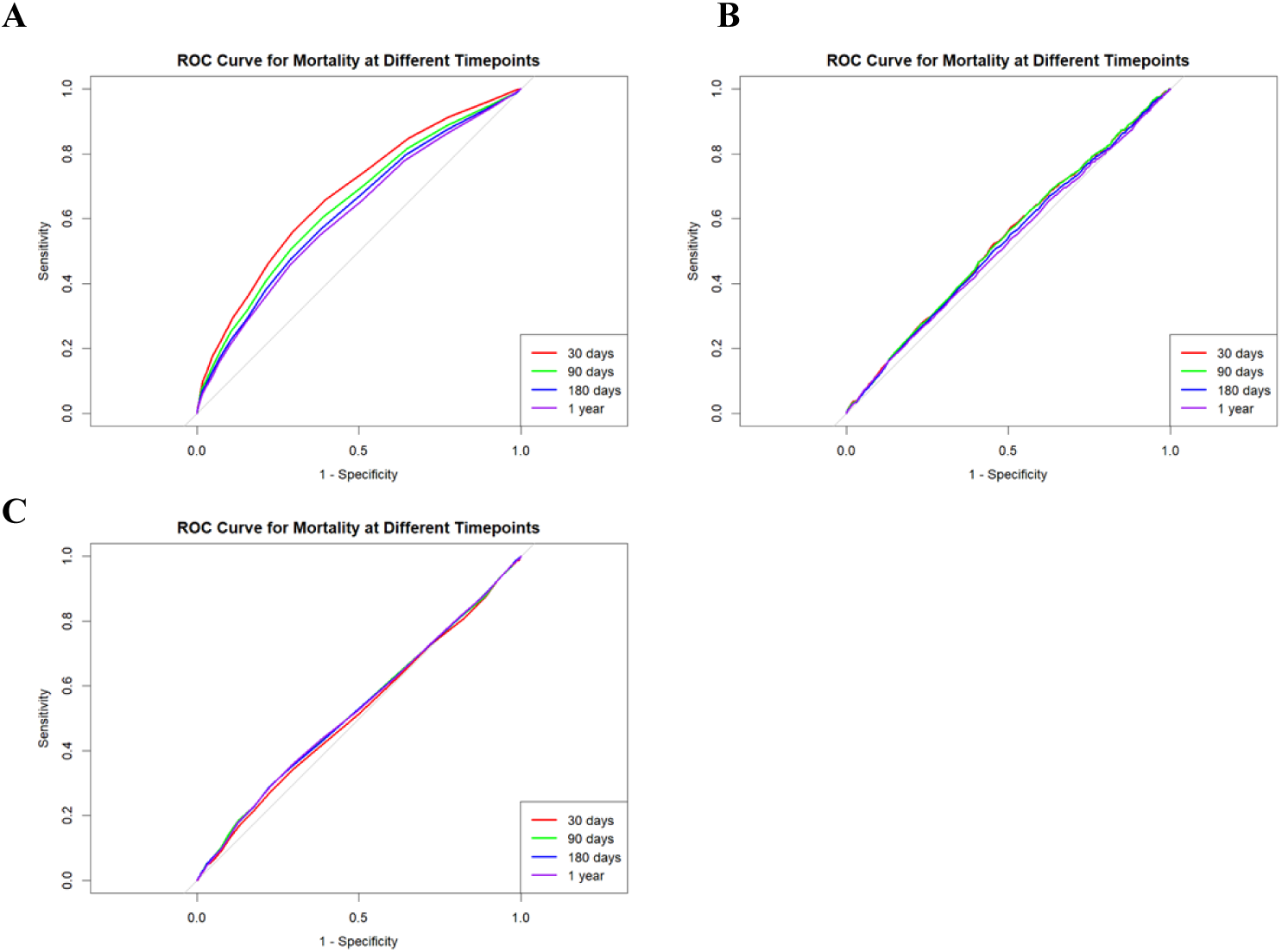
ROC curves for mortality at different time points. Note: A, B, and C are ROC curves for GPR, glucose, and potassium at 28 days, 90 days, 180 days, and 1 year, respectively.

## Discussion

In this study, we found that the serum glucose-potassium ratio (GPR) is an independent predictor of all-cause mortality (ACM) in sepsis patients. Higher GPR values were significantly associated with lower mortality risk at multiple time points, including 28 days, 90 days, 180 days, and 1 year. This finding suggests that GPR may provide important prognostic information for clinical risk assessment in sepsis.

Early recognition and timely intervention in sepsis are crucial for improving prognosis. However, due to the complexity and diverse clinical presentations of sepsis, traditional clinical scoring systems (e.g., SOFA, APACHE II scores) often fail to accurately assess mortality risk in these patients[6]. By analyzing data from the MIMIC-IV database, we found that GPR offers a more sensitive prognostic prediction. Compared to glucose or potassium levels alone, GPR simultaneously reflects a patient’s glucose metabolism status and electrolyte balance[18], which are particularly important in sepsis, where stress-induced hyperglycemia and electrolyte disturbances are commonly observed[19].

High GPR values may indicate more severe metabolic disturbances, which are closely linked to organ dysfunction associated with sepsis. For example, hyperglycemia is associated with immune dysfunction, suppressed inflammatory responses, and worsening sepsis outcomes[20,21]. Additionally, hypokalemia can exacerbate cardiovascular instability, contributing to further organ dysfunction[22].

The biological significance of GPR, as a composite ratio of blood glucose to potassium, lies in the synergistic roles of these two factors in sepsis pathophysiology. Blood glucose and potassium levels are intricately linked through mechanisms mediated by insulin and catecholamines (e.g., epinephrine). These hormones activate β2-adrenergic and insulin receptors on cell surfaces, enhancing Na+-K+ ATPase pump activity. This process facilitates potassium influx into cells while promoting glucose uptake, thereby lowering both serum potassium and glucose levels[23,24].

Elevated blood glucose stimulates insulin secretion, which drives potassium into cells, reducing serum potassium levels. Conversely, high glucose levels can further influence potassium homeostasis through insulin-mediated mechanisms, creating a dynamic interplay between these metabolic parameters[23,24]. GPR reflects the body’s metabolic stress state, often associated with endocrine responses and neuroendocrine system activation[24]. During acute stress, such as infection or trauma, sympathetic nervous system activation typically elevates blood glucose while reducing potassium levels, disrupting cellular metabolism and electrolyte balance.

GPR, as an indicator of metabolic disturbances, reflects dysregulation of glucose metabolism and electrolyte imbalance in sepsis patients. In critically ill populations, a higher GPR is typically associated with worse clinical outcomes. These metabolic disturbances can exacerbate the systemic inflammatory response and impair organ function[25]. For instance, an elevated GPR often signifies severe metabolic stress and endocrine dysregulation, increasing the risk of multi-organ failure and mortality[18].

This study has limitations: (1) potential selection bias and unmeasured confounders due to its single-center, retrospective design (MIMIC-IV database); (2) lack of dynamic GPR monitoring during treatment; and (3) insufficient exploration of molecular mechanisms linking GPR to sepsis pathophysiology (e.g., oxidative stress, coagulation).

Future research should: (1) validate GPR’s predictive efficacy in multicenter prospective studies and develop integrated models (e.g., GPR-SOFA); (2) explore molecular mechanisms (e.g., insulin signaling, immune cell polarization) through experimental models; (3) conduct clinical trials (e.g., potassium supplementation with glycemic control) for low-GPR patients; and (4) expand studies in diverse populations to analyze genetic influences.

## Conclusion

GPR is a robust, independent predictor of sepsis mortality, outperforming traditional markers, particularly in elderly, female, and Caucasian populations. Despite the retrospective nature of the study, its threshold stability and clinical utility support its potential as a bedside tool. Further mechanistic and interventional studies are needed to validate its clinical application.

## Data Availability

All relevant data are within the manuscript and its Supporting Information files.

## Acknowledgments

We would like to express our sincere gratitude to all those who contributed to this study. First, we thank the MIMIC-IV database team for providing access to the valuable data used in this research. Their work is indispensable for advancing healthcare research through the availability of high-quality clinical data. Finally, we would like to express our gratitude to the anonymous reviewers and editors of this journal for their invaluable feedback and suggestions, which greatly enhanced the clarity and rigor of this manuscript.

## Notes

### Competing Interest Statement

The authors have declared no competing interest.

### Funding Statement

The author(s) received no specific funding for this work.

### Author Declarations

Massachusetts Institute of Technology and Beth Israel Deaconess Medical Center

